# Deficits in hospital care among clinically vulnerable children aged 0 to 4 years during the COVID-19 pandemic

**DOI:** 10.1101/2021.12.16.21267904

**Authors:** David Etoori, Katie Harron, Louise Mc Grath-Lone, Maximiliane Verfuerden, Ruth Gilbert, Ruth Blackburn

## Abstract

**Objective:** To quantify deficits in hospital care for clinically vulnerable children during the COVID-19 pandemic.

**Design:** Birth cohort in Hospital Episode Statistics (HES).

**Setting:** NHS hospitals in England.

**Study population:** All children aged <5 years with a birth recorded in hospital administrative data (January 2010 to March 2021).

**Main exposure:** Clinical vulnerability defined by a chronic health condition, preterm birth (<37 weeks gestation) or low birthweight (<2500g).

**Main outcomes:** Deficits in care defined by predicted rates for 2020, estimated from 2015- 2019, minus observed rates per 1000 child years during the pandemic (March 2020-2021).

**Results:** Of 3,813,465 children, 17.7% (1 in 6) were clinically vulnerable (9.5% born preterm or low birthweight, 10.3% had a chronic condition). Deficits in hospital care during the pandemic were much higher for clinically vulnerable children than peers: respectively, outpatient attendances (314 versus 73 per 1000 child years), planned admissions (55 versus 10), and unplanned admissions (105 versus 79). Clinically vulnerable children accounted for 50.1% of the deficit in outpatient attendances, 55.0% in planned admissions, and 32.8% in unplanned hospital admissions. During the pandemic, weekly rates of planned care returned to pre-pandemic levels for infants with chronic conditions but not older children. Deficits in care differed by ethnic group and level of deprivation. Virtual outpatient attendances increased from 3.2% to 24.8% during the pandemic.

**Conclusion:** 1 in 6 clinically vulnerable children accounted for one-third to one half of the deficit in hospital care during the pandemic.

## Introduction

Rates of hospital contact among 0-4 year olds increased steadily in England over the past decade and are highest for infants.(1–3) Hospital utilisation patterns differ markedly by age and clinical vulnerability: children born preterm (<37 weeks of gestation), with low birth weight (<2500g) or a congenital anomaly, have substantially more admissions than other children.(4,5)

Hospital contacts declined substantially during the COVID lockdown.(6–9) This decline is likely to have impacted most on children with high rates of planned hospital care, such as those born too early or too small or with underlying health conditions.(1,2,10) Planned hospital care that was postponed or cancelled may result in delayed diagnoses or treatments, which could be detrimental to health or development.(11) Fewer unplanned hospital admissions might reflect fewer infections, injuries, or other health problems due to reduced exposure during pandemic restrictions, but could also reflect unmet need for unplanned care.

This study aimed to quantify deficits in planned and unplanned hospital care for clinically vulnerable children and their peers during the COVID-19 pandemic. We used national, longitudinal administrative data for England to measure rates of planned hospital contacts (admissions and outpatient attendances) and unplanned admissions among children with chronic health conditions or born too early or too small, and their peers. We quantified the deficit by comparing predicted and observed rates of hospital contacts during the first year of the pandemic. We examined whether rates of contact returned to pre-pandemic levels and described changes in the type of outpatient contact (e.g., face-to-face or virtual).

## Methods

### Study population and data source

Children were included in the cohort if aged 0-4 years and their birth was recorded in hospital administrative data in the English NHS (Hospital Episode Statistics – HES) between January 1, 2010, and March 31, 2021. Births recorded in HES represent 97% of all births in England.(12) Children were followed until their fifth birthday or March 31, 2021, whichever occurred first (Supplementary Figure 1). All contacts with NHS hospitals in England were routinely linked, including admitted patient care (APC) and outpatient attendances. Due to changes in linkage methods used by NHS Digital, accident and emergency (A&E) attendances could not be included.(13) We combined consecutive consultant episodes and hospital transfers to form admissions.(14)

### Outcome and exposure

Our primary outcome was the deficit in hospital care, defined as the absolute difference between observed and predicted rates (per 1,000 child-years) of hospital contacts (outpatient attendances, planned and unplanned hospital admissions) during the pandemic (March 23, 2020, to March 22, 2021). We also described trends in weekly rates of hospital contacts by year of age (from January 1, 2020 to March 31, 2021, and averaged for 2015-2019), and uptake (attended, missed, cancelled, postponed) and mode (in person, virtually) of outpatient contacts.

Clinical vulnerability was defined by a chronic condition recorded in HES up to age 4, or preterm birth (<37 weeks of gestation) or low birth weight (<2500g) recorded in birth or delivery records, using previously reported methods.(12,15–17) Children with missing gestation and birth weight data who had no chronic conditions were categorised as having no clinical vulnerability.

We analysed the following risk factors associated with frequency of hospital contacts: age (0 to 11 months, 1 to 4 years),(3) quintile of deprivation derived from the national distribution of the Index of Multiple Deprivation (IMD) 2004 (an area measure for ∼650 households),(18,19) and ethnic group recorded in HES (grouped as White, Black, Asian, or Other, including mixed and Chinese).

### Statistical analyses

We calculated observed rates of hospital contacts per 1,000 child-years in the pre-pandemic period (2015-2019), stratifying by risk factor (clinical vulnerability, age group, area quintile of deprivation, and ethnic group).

We calculated child-years at risk by averaging the eligible population of births recorded in HES at the beginning and end of a year, assuming no emigration and ignoring deaths. We used a Poisson model, which included a linear effect of time and log of the mid-year population as an offset, to model rates from January 1, 2015, to December 31, 2019 stratifying by risk factors. Data from January 1 to March 22, 2020, was excluded from the pre-pandemic period as we observed reductions in hospital contact rates before the first lockdown. To calculate the deficit in hospital contacts, we predicted rates for the pandemic period, assuming that the pandemic had not occurred and that previous trends would have continued. The deficit was estimated as the difference between predicted and observed rates. We also calculated deficits within the first national lockdown (March 23 to June 23 2020), easing of restrictions (June 24 to November 4 2020), second national lockdown (November 5 – December 31 2020), and third national lockdown (January 1 – March 22 2021).(20)

To understand seasonality and visualise the decline and recovery in hospital contacts during the pandemic, we calculated weekly rates between January 1, 2015, and March 31, 2021 using dynamic denominator study populations. Since we expected the largest differences to be seen when comparing children with and without chronic conditions, we stratified by this risk factor as recorded in any HES admission between birth and the relevant week. Weekly rates of hospital contact were calculated by dividing the total number of weekly admissions or attendances by the weekly dynamic denominator population of children within each stratification level (i.e., a child born in week 1 of 2015 would move into the 1-year-old group in week 1 of 2016 and age out of the cohort in week 1 of 2019). We then visually compared weekly plots of rates in 2020 and 2021 with the overall average rates for 2015-2019 for the same week. We also modelled weekly rates between January 1, 2015, and December 31, 2019, using a Poisson model which included a linear effect of time, calendar month to account for seasonality, log of the weekly denominator population as an offset, and second order lagged residuals. A similar approach was then used to estimate weekly deficits during the pandemic. All analyses were performed in Stata 16.(21)

## Results

### Population characteristics

Of the 3,813,465 children aged 0-4 years, 394,384 (10.3%) had a record indicating a chronic condition (including congenital anomalies); 363,950 (9.5%) were born preterm or low birthweight, and 83,283 (2.2%) had both vulnerabilities. Overall, 675,051 (17.7%) had one or more of these clinical vulnerabilities (Table 1).

**Table 1:** Demographic characteristics of children born between January 1, 2015, and March 31, 2021, by vulnerability status.

|  |  | None<br>(N=3,138,414; 82.3%) |  | LBW/Preterm only<br>(N=280,667; 7.4%) |  | CC only<br>(N=311,101; 8.2%) |  | Both<br>(N=83,283; 2.2%) |  | Total<br>(N=3,813,465) |  |
| --- | --- | --- | --- | --- | --- | --- | --- | --- | --- | --- | --- |
|  |  | n | % | n | % | n | % | n | % | n | % |
| **Sex** | Male | 1,590,308 | 50.7 | 134,871 | 48.1 | 183,964 | 59.1 | 46,304 | 55.6 | 1,955,447 | 51.3 |
|  | Female | 1,547,082 | 49.3 | 145,481 | 51.8 | 127,006 | 40.8 | 36,860 | 44.3 | 1,856,429 | 48.7 |
|  | Missing | 1,024 | 0.03 | 315 | 0.1 | 131 | 0.04 | 119 | 0.1 | 1,589 | 0.04 |
| **Age group** | Infants | 480,832 | 15.3 | 36,445 | 13.0 | 27,641 | 8.9 | 8,317 | 10.0 | 553,235 | 14.5 |
|  | 1-4-year-olds | 2,657,582 | 84.7 | 244,222 | 87.0 | 283,460 | 91.1 | 74,966 | 90.0 | 3,260,230 | 85.5 |
| **Ethnic group** | White | 2,185,934 | 69.7 | 184,525 | 65.8 | 228,023 | 73.3 | 58,366 | 70.1 | 2,656,848 | 69.7 |
|  | Black/Black British | 140,991 | 4.5 | 15,477 | 5.5 | 14,150 | 4.6 | 4,681 | 5.6 | 175,299 | 4.6 |
|  | Asian/Asian British | 332,009 | 10.6 | 41,516 | 14.8 | 33,610 | 10.8 | 10,857 | 13.0 | 417,992 | 11.0 |
|  | Any other ethnic groups | 273,658 | 8.7 | 24,085 | 8.6 | 25,135 | 8.1 | 6,633 | 8.0 | 329,511 | 8.6 |
|  | Missing | 205,822 | 6.6 | 15,064 | 5.4 | 10,183 | 3.3 | 2,746 | 3.3 | 233,815 | 6.1 |
| **Deprivation quintile** | Q1 (most deprived) | 739,549 | 23.6 | 83,204 | 29.7 | 89,425 | 28.7 | 27,254 | 32.7 | 939,432 | 24.6 |
|  | Q2 | 612,665 | 19.5 | 60,337 | 21.5 | 67,853 | 21.8 | 18,629 | 22.4 | 759,484 | 19.9 |
|  | Q3 | 524,863 | 16.7 | 45,794 | 16.3 | 55,924 | 18.0 | 14,117 | 17.0 | 640,698 | 16.8 |
|  | Q4 | 461,118 | 14.7 | 37,446 | 13.3 | 47,670 | 15.3 | 11,287 | 13.6 | 557,521 | 14.6 |
|  | Q5 (least deprived) | 436,147 | 13.9 | 34,530 | 12.3 | 43,052 | 13.8 | 10,119 | 12.2 | 523,848 | 13.7 |
|  | Missing | 364,072 | 11.6 | 19,356 | 6.9 | 7,177 | 2.3 | 1,877 | 2.3 | 392,482 | 10.3 |
| **Year of birth** | 2015 | 512,624 | 16.3 | 46,590 | 16.6 | 68,814 | 22.1 | 16,021 | 19.2 | 644,049 | 16.9 |
|  | 2016 | 515,510 | 16.4 | 49,020 | 17.5 | 63,505 | 20.4 | 16,042 | 19.3 | 644,077 | 16.9 |
|  | 2017 | 511,838 | 16.3 | 49,358 | 17.6 | 55,396 | 17.8 | 15,096 | 18.1 | 631,688 | 16.6 |
|  | 2018 | 500,133 | 15.9 | 47,406 | 16.9 | 47,628 | 15.3 | 13,730 | 16.5 | 608,897 | 16.0 |
|  | 2019 | 497,682 | 15.9 | 41,713 | 14.9 | 40,107 | 12.9 | 11,646 | 14.0 | 591,148 | 15.5 |
|  | 2020 | 487,365 | 15.5 | 38,128 | 13.6 | 30,598 | 9.8 | 9,341 | 11.2 | 565,432 | 14.8 |
|  | 2021 | 113,262 | 3.6 | 8,452 | 3.0 | 5,053 | 1.6 | 1,407 | 1.7 | 128,174 | 3.4 |

**LBW** – Low birthweight; **CC** – Chronic conditions

### Hospital contacts pre-pandemic

Hospital contacts were much higher among infants than children aged 1-4 years: 60.1% of infants and 8.2% of 1–4-year-olds had ≥1 outpatient attendance each year (Supplementary Figure 2, Supplementary Table 1). Overall, 31.2% of clinically vulnerable children had ≥1 outpatient attendance compared with 14.1% of those with no known vulnerability (p<0.001). A similar pattern was seen for planned and unplanned hospital admissions. Children with chronic conditions had the highest rates of planned and unplanned admissions across all strata (Supplementary Figures 3-4). In contrast, children born preterm or low birth weight but with no chronic condition had similar rates of admissions to their peers born at term or weighing ≥2500g (Supplementary Figure 2).

### Hospital contacts during the pandemic

There were stark reductions in rates of all types of hospital care during the pandemic (Table 2, Supplementary tables 2-4). Differences between predicted and observed rates of hospital contacts were much larger for children with a chronic condition than those without, and particularly high for children with a chronic condition who were also born preterm or with a low birth weight (Table 2). The 17.7% of children who were categorised as clinically vulnerable accounted for 50.1% of the deficit in outpatient attendances, 55.0% of the deficit in planned hospital admissions, and 32.8% of the deficit in unplanned hospital admissions (Table 2).

**Table 2:** Difference between predicted and observed rates of hospital contact during the pandemic (March 2020-2021) among children aged 0 to 4 years, by clinical vulnerability group.

|  | Percentage of children seen |  |  | Number of hospital contacts |  |  |  | Rates per 1000 child-years |  |  |  |
| --- | --- | --- | --- | --- | --- | --- | --- | --- | --- | --- | --- |
| Outpatient attendances | n | N | % | Predicted | Observed | Difference | % change | Predicted | Observed | Difference (95% CI) | % change |
| Total | 370,623 | 2,765,941 | 13.4 | 1,500,674 | 1,173,091 | -327,583 | -21.8 | 543 | 424 | **-118** (-115, -122) | -21.8 |
| No known vulnerability | 232,976 | 2,243,088 | 10.4 | 635,226 | 471,743 | -163,483 | -25.7 | 283 | 210 | **-73** (-71, -74) | -25.7 |
| Any vulnerability | 137,647 | 522,853 | 26.3 | 865,448 | 701,348 | -164,100 | -19.0 | 1655 | 1341 | **-314\*** (-302, -326) | -19.0 |
| • LBW/Preterm only | 29,622 | 226,439 | 13.1 | 94,660 | 76,039 | -18,621 | -19.7 | 418 | 336 | **-82\*** (-77, -88) | -19.7 |
| • CC only | 86,825 | 231,671 | 37.5 | 586,026 | 475,226 | -110,800 | -18.9 | 2530 | 2051 | **-478\*** (-465, -492) | -18.9 |
| • Both | 21,200 | 64,743 | 32.7 | 184,762 | 150,083 | -34,679 | -18.8 | 2854 | 2318 | **-536\*** (-508, -563) | -18.8 |
| Any CC<sup>α</sup> | 107,762 | 296,414 | 36.4 | 771,219 | 625,309 | -145,910 | -18.9 | 2602 | 2110 | **-492\*** (-480, -505) | -18.9 |
| Planned admissions |  |  |  |  |  |  |  |  |  |  |  |
| Total | 52,280 | 2,765,941 | 1.9 | 144,980 | 92,618 | -52,362 | -36.1 | 52 | 33 | **-19** (-18, -20) | -36.1 |
| No known vulnerability | 15,287 | 2,243,088 | 0.7 | 40,568 | 17,022 | -23,546 | -58.0 | 18 | 8 | **-10** (-10, -11) | -58.0 |
| Any vulnerability | 36,993 | 522,853 | 7.1 | 104,412 | 75,596 | -28,816 | -27.6 | 200 | 145 | **-55\*** (-51, -59) | -27.6 |
| • LBW/Preterm only | 1,389 | 226,439 | 0.6 | 3,675 | 1,522 | -2,153 | -58.6 | 16 | 7 | **-10** (-8, -11) | -58.6 |
| • CC only | 31,038 | 231,671 | 13.4 | 85,367 | 65,506 | -19,861 | -23.3 | 368 | 283 | **-86\*** (-81, -91) | -23.3 |
| • Both | 4,566 | 64,743 | 7.1 | 15,370 | 8,568 | -6,802 | -44.3 | 237 | 132 | **-105\*** (-97, -113) | -44.3 |
| Any CC | 35,603 | 296,414 | 12.0 | 100,929 | 74,074 | -26,855 | -26.6 | 341 | 250 | **-91\*** (-86, -95) | -26.6 |
| Unplanned admissions |  |  |  |  |  |  |  |  |  |  |  |
| Total | 131,134 | 2,765,941 | 4.7 | 442,174 | 179,366 | -262,808 | -59.4 | 160 | 65 | **-95** (-93, -97) | -59.4 |
| No known vulnerability | 66,830 | 2,243,088 | 3.0 | 254,263 | 77,713 | -176,550 | -69.4 | 113 | 35 | **-79** (-78, -80) | -69.4 |
| Any vulnerability | 64,304 | 522,853 | 12.3 | 187,911 | 101,653 | -86,258 | -45.9 | 359 | 194 | **-165\*** (-159, -171) | -45.9 |
| • LBW/Preterm only | 6,046 | 226,439 | 2.7 | 25,520 | 7,228 | -18,292 | -71.7 | 113 | 32 | **-81** (-78, -84) | -71.7 |
| • CC only | 52,285 | 231,671 | 22.6 | 134,055 | 84,126 | -49,929 | -37.2 | 579 | 363 | **-216\*** (-209, -222) | -37.2 |
| • Both | 5,973 | 64,743 | 9.2 | 28,336 | 10,299 | -18,037 | -63.7 | 438 | 159 | **-279\*** (-268, -290) | -63.7 |
| Any CC | 58,248 | 296,414 | 19.7 | 162,648 | 94,425 | -68,223 | -41.9 | 549 | 319 | **-230\*** (-224, -236) | -41.9 |

**LBW** – Low birthweight; **CC** – Chronic conditions. <sup>α</sup> Any CC combines CC only and both. \* Significantly different from children with no known vulnerability (p<0.05).

Deficits were higher for infants than 1-4-year-olds for outpatient attendances and unplanned admissions but not for planned admissions (Figure 1). We found small but statistically significant differences in deficits of planned and unplanned admissions across ethnic groups and in all hospital contacts among children in the most (versus least) deprived quintile (Supplementary tables 3-4). The largest deficits in care was among children with a chronic condition (Supplementary Figures 5-6). Overall, the largest deficits were seen in the first lockdown for outpatient attendances and planned admissions, but for unplanned admissions, the largest deficits occurred in the second lockdown period (Table 3; Supplementary tables 5-7).

**Figure 1:**
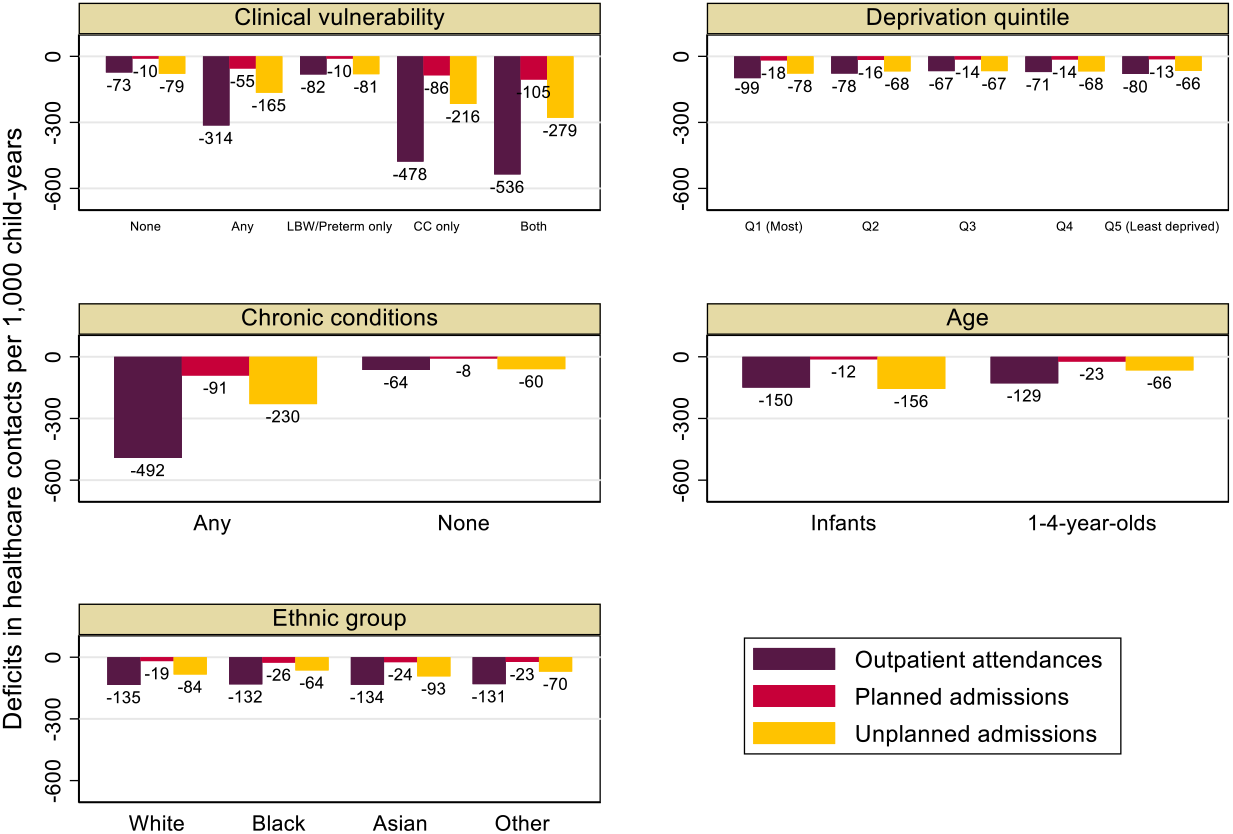
Deficit in care during the pandemic (March 2020-2021), estimated from predicted minus observed rates of hospital contacts per 1,000 child-years for children aged 0 to 4 years, by clinical vulnerability status and risk factors. **LBW** – Low birthweight; **CC** – Chronic conditions

**Table 3:** Difference in predicted and observed rates of hospital contact per 1,000 child-years among children aged 0 to 4 years during the pandemic (March 2020-2021), by period and clinical vulnerability.

|  | 1<sup>st</sup> lockdown<br>March 23 to June 23, 2020 |  | Easing of restrictions<br>June 24 to Nov 4, 2020 |  | 2<sup>nd</sup> lockdown<br>Nov 5 to Dec 31, 2020 |  | 3<sup>rd</sup> lockdown<br>Jan 1 to March 22, 2021 |  |
| --- | --- | --- | --- | --- | --- | --- | --- | --- |
| **Outpatient attendances** | Deficit (95% CI) | % change | Deficit (95% CI) | % change | Deficit (95% CI) | % change | Deficit (95% CI) | % change |
| **Total** | **-167** (-163, -170) | -30.7 | **-88** (-86, -91) | -16.3 | **-96** (-91, -100) | -17.9 | **-128** (-123, -132) | -23.5 |
| No known vulnerability | **-106** (-104, -107) | -37.2 | **-55** (-54, -56) | -19.5 | **-56** (-55, -58) | -20.6 | **-75** (-73, -77) | -26.2 |
| Any vulnerability | **-428\*** (-417, -439) | -25.9 | **-230\*** (-221, -240) | -13.9 | **-264\*** (-249, -278) | -16.0 | **-352\*** (-337, -367) | -21.4 |
| • LBW/Preterm only | **-121\*** (-116, -127) | -28.7 | **-57** (-53, -61) | -13.8 | **-73\*** (-67, -80) | -18.0 | **-84** (-77, -92) | -19.9 |
| • CC only | **-664\*** (-651, -677) | -26.2 | **-357\*** (-346, -368) | -14.0 | **-398\*** (-381, -414) | -15.9 | **-516\*** (-499, -533) | -20.8 |
| • Both | **-655\*** (-629, -681) | -23.3 | **-385\*** (-364, -407) | -13.7 | **-450\*** (-416, -483) | -15.5 | **-699\*** (-665, -735) | -24.2 |
| Any CC | **-663\*** (-652, -675) | -25.5 | **-364\*** (-354, -374) | -14.0 | **-410\*** (-395, -425) | -15.8 | **-559\*** (-544, -574) | -21.7 |
| **Planned admissions** |  |  |  |  |  |  |  |  |
| **Total** | **-32** (-31, -33) | -59.2 | **-16** (-15, -17) | -29.3 | **-9** (-8, -11) | -19.2 | **-16** (-15, -17) | -31.9 |
| No known vulnerability | **-14** (-14, -15) | -79.1 | **-9** (-9, -9) | -48.5 | **-7** (-6, -7) | -38.9 | **-11** (-11, -12) | -62.0 |
| Any vulnerability | **-105\*** (-101, -109) | -51.6 | **-46\*** (-43, -49) | -22.0 | **-22\*** (-18, -26) | -11.6 | **-36\*** (-32, -41) | -19.4 |
| • LBW/Preterm only | **-12\*** (-11, -13) | -75.3 | **-8** (-7, -9) | -49.4 | **-6** (-5, -7) | -42.1 | **-11** (-11, -10) | -64.9 |
| • CC only | **-185\*** (-180, -190) | -49.0 | **-71\*** (-67, -75) | -18.4 | **-24\*** (-18, -30) | -6.9 | **-40\*** (-34, -46) | -11.6 |
| • Both | **-144\*** (-137, -152) | -60.3 | **-86\*** (-80, -93) | -35.8 | **-70\*** (-61, -79) | -31.6 | **-114\*** (-105, -125) | -48.2 |
| Any CC | **-177\*** (-172, -181) | -50.8 | **-75\*** (-71, -78) | -21.2 | **-35\*** (-30, -40) | -10.7 | **-57\*** (-52, -62) | -17.7 |
| **Unplanned admissions** |  |  |  |  |  |  |  |  |
| **Total** | **-88** (-87, -90) | -60.0 | **-77** (-76, -79) | -51.9 | **-140** (-138, -143) | -66.2 | **-99** (-97, -101) | -64.2 |
| No known vulnerability | **-68** (-67, -69) | -66.5 | **-66** (-66, -67) | -63.2 | **-116** (-115, -118) | -75.3 | **-84** (-83, -85) | -76.7 |
| Any vulnerability | **-176\*** (-171, -182) | -51.6 | **-124\*** (-120, -128) | -36.9 | **-245\*** (-237, -253) | -53.2 | **-161\*** (-154, -168) | -47.1 |
| • LBW/Preterm only | **-69** (-66, -72) | -68.0 | **-64** (-62, -66) | -64.1 | **-125\*** (-121, -130) | -78.3 | **-90\*** (-86, -94) | -79.9 |
| • CC only | **-263\*** (-257, -270) | -47.2 | **-157\*** (-152, -162) | -28.5 | **-309\*** (-300, -318) | -43.1 | **-189\*** (-181, -197) | -34.8 |
| • Both | **-241\*** (-231, -251) | -59.1 | **-216\*** (-208, -225) | -54.8 | **-434\*** (-419, -450) | -73.1 | **-311\*** (-298, -325) | -72.8 |
| Any CC | **-259\*** (-254, -265) | -49.3 | **-171\*** (-166, -175) | -33.0 | **-337\*** (-330, -345) | -48.8 | **-217\*** (-210, -224) | -41.8 |

**LBW** – Low birthweight; **CC** – Chronic conditions \* Significantly different from children with no known vulnerability (5% level of significance).

### Trends in hospital contacts

Outpatient attendances declined sharply before and during the first national lockdown (Figure 2), among children of all ages with a chronic condition, with less perceptible changes among those without a chronic condition. Outpatient attendances rapidly returned to pre-pandemic rates for infants, but not for older children which remained below 2015-19 averages in the third lockdown. Planned admissions followed a similar pattern, with a return to pre-pandemic rates only for infants (Figure 2).

**Figure 2:**
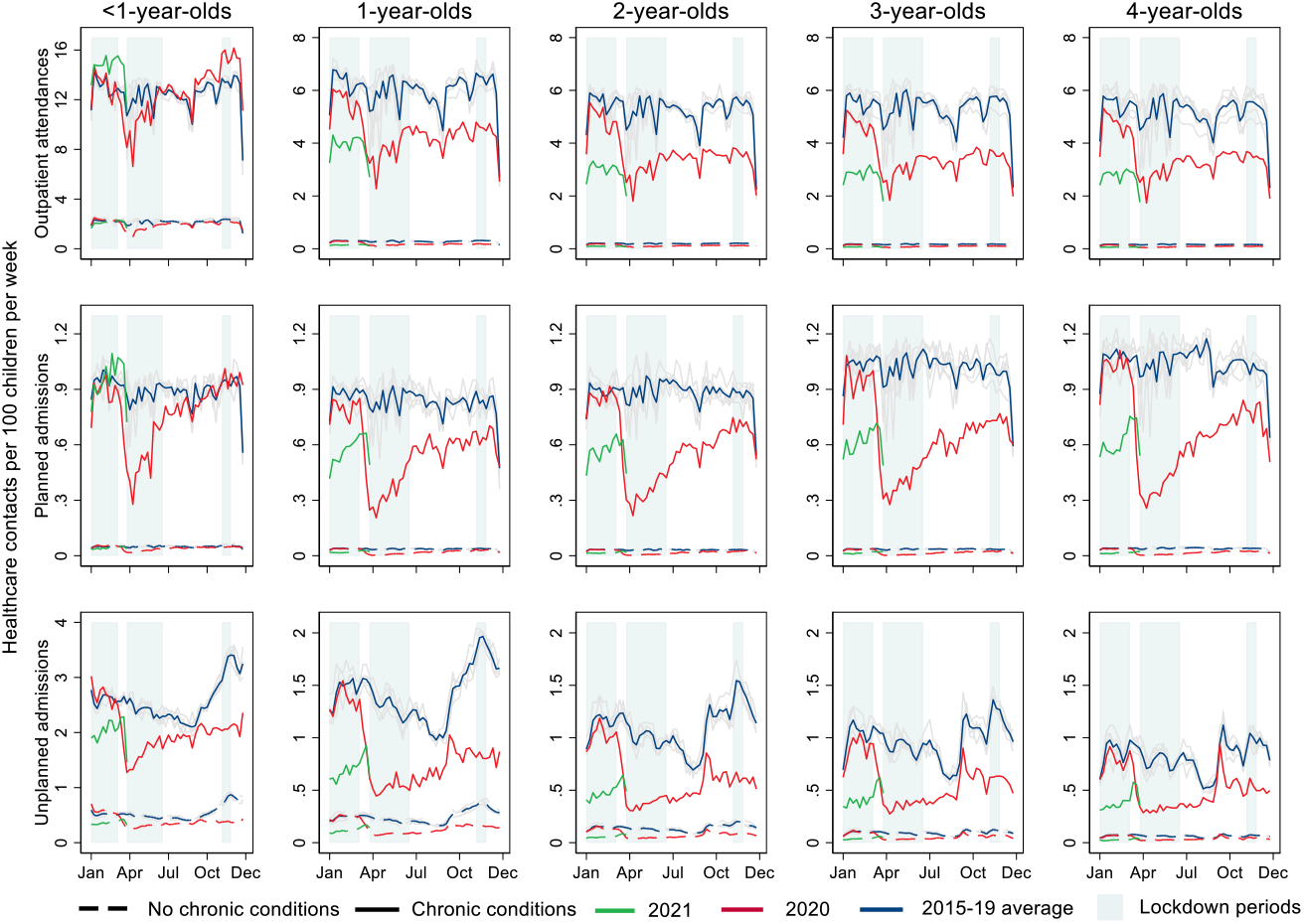
Weekly rates of hospital contacts among children aged 0 to 4 years during the pandemic (March 2020-2021) and averaged for 2015-2019, by age and presence of a chronic condition. Note: January-March lockdown only affects 2021 data.

Declines in rates of unplanned admissions during the first lockdown were much greater for children with a chronic condition than those without and remained below pre-pandemic levels for both groups at all ages (Figure 2). In 2020, the autumn-winter peak in unplanned admissions was diminished relative to previous years; however, following the reopening of primary schools at the end of the third lockdown on March 8, 2021, there was an increase in unplanned admission rates for all children, particularly those with a chronic condition (Figure 2, Supplementary Figure 7). Trends did not differ consistently according to deprivation level (Supplementary Figures 8-9) or by ethnic group (Supplementary Figures 10-12).

For all age groups, a spike in cancellations and postponement of outpatient appointments preceded the first lockdown by three weeks (Supplementary Figure 13). There was an increase in tele/virtual outpatient attendances during the pandemic and face-to-face visits did not return to pre-pandemic levels in any age group (Supplementary Table 8 and Figure 14).

## Discussion

This population-based cohort study of all children aged <5 in England found large and disproportionate deficits in planned and unplanned hospital contacts during the COVID-19 pandemic for clinically vulnerable groups and those living in deprived areas. The one in six clinically vulnerable children accounted for over half the deficit in outpatient attendances and planned admissions, and one-third of the deficit in unplanned admissions. We saw some evidence of recovery in the deficit in planned care during the pandemic among infants, but not among older children.

This study has several strengths. First, we used a birth cohort of all children born in an NHS hospital in England (97% of all births). Our rates did not account for deaths in the denominator (0.5%), non-NHS health care, or emigration, but these events are rare. Second, HES captures gestational age and birthweight and we used a clinically developed coding system to define chronic conditions from diagnostic codes recorded in admissions.(15) Third, the large study population made it possible to calculate weekly rates. Fourth, we used data preceding the pandemic to March 2021 to investigate variation across the first year of the pandemic.

Limitations include under-ascertainment of chronic conditions for children who could not be admitted to hospital due to the pandemic. These children may have been managed in primary care, or as outpatients, which does not reliably code chronic conditions. Our analyses may underestimate vulnerability for the 10% of children without a record of gestational age or birthweight as we assumed they had non-vulnerable status. Multiple imputation of missing data was not feasible given the study size. Second, we could not quantify the deficit in A&E attendances, as longitudinal linkage is not currently available. However, studies investigating A&E attendances have reported similar deficits to those found in our study.(22,23) Our modelling approach required several assumptions (including continued trends in hospital contacts), and the differences we report are likely conservative estimates of impact.

Deficits in hospital care for children during the pandemic have been reported in Europe,(7,24–31) Asia,(32) North,(33–36) and South America.(37) Most studies investigated A&E attendances or unplanned admissions.(23–37) Other studies report a reduction in asthma-related paediatric emergency department attendances,(27) and reduced likelihood of admission, assessment and surgery for children with epilepsy.(38) Furthermore, significant reductions in infection-related hospitalisations have been observed,(28,32,34,39) particularly for children under 5 years.(39) Two studies conducted national level analyses.(28,30) We believe our study is the first to report on deficits in planned care (admissions and outpatient), on disproportionate deficits in hospital care for children with health risk-factors at a population-level, and for a full year of the pandemic. Previous research in adult populations has reported on the disproportionate burden of COVID-19 infection, hospitalisation, and death in minority ethnic groups.(40,41) Our study did not examine COVID-related contacts because children hospitalisation is rare as typically experience mild asymptomatic disease.(42,43) However, we identified small differences in deficits of hospital care for children in the Asian ethnic group, and for children in the most deprived quintile, indicating evidence of structural inequalities in healthcare access.

Potential mechanisms underpinning deficits in planned care likely represent restrictions to access, supported by a rise in postponements of outpatient care. Our findings show these restrictions were mitigated in infants, who have a high frequency of hospital care and for whom interventions are likely to be more time-critical than in older children.(3) However, deficits remained large in older children and may reflect unmet need or postponed care that could have longer-term health and care consequences.(11) Furthermore, children without access to the internet at home will have been disproportionately impacted by the move to virtual appointments.

Deficits in unplanned care may be driven by opposing factors. Previous studies reported reductions in unplanned admissions due to decreases in infection-related hospitalisations, due to reduced social exposure and increased hygiene, with little change in admissions for non-infectious causes such as appendicitis.(25,28,29,32,39) Others have reported reductions in injury.(44) The spike in unplanned admissions after schools reopened in autumn 2020 and in March 2021 when the third lockdown ended likely reflects increased socialisation. Other positive effects could include reduced exposure to triggers for respiratory disease (e.g., air pollution),(45) improved hygiene due to ubiquitous handwashing messaging, and improved medication adherence through increased parental supervision. Negative implications could include reduced extrinsic interventions through education, health, and social care professionals,(7,46) or delaying or avoiding medical care due to restricted access or fears of potential hospital-acquired COVID-19 infection.(36,47– 49) Additionally, these deficits could represent a missed opportunity to catch problems early which could also have consequences in the future.(11,50)

This analysis was the first step in quantifying the burden of deferred or foregone hospital care during the pandemic and the potential consequences for young children. Studies using routine administrative data report only the most acute cases are presenting at hospitals,(35,36) and that some of the deficits reflect late or missed diagnoses.(11,50) More research is needed to untangle these different mechanisms. Research is also needed to understand deficits in planned care, the types of care, procedures, or treatments affected and the short-and long-term implications for children with specific conditions. Our findings indicate a need for targeted ‘catch-up’ funding and resources for child health, particularly for vulnerable children who were affected disproportionately.(51)

## Supporting information

STROBE checklist

## Data Availability

The data used in this analysis is expected to be available to accredited researchers in 2022 (as part of the ECHILD Database) by applying to the data providers (Department for Education and NHS Digital).

## Acknowledgements

The data for this project is part of the ECHILD project. The ECHILD project is in partnership with NHS Digital and the Department for Education (DfE) and we thank the following individuals for their valuable contributions to the project: Garry Coleman, Richard Caulton, Joanna Geisler, Catherine Day (NHS Digital), Chris Douglass and Gary Connell (DfE).

We thank all the children, young people, parents and carers who contributed to the ECHILD project, as well as Dr Erin Walker (UCL Partners) who led this involvement. We would particularly like to thank members of the National Children’ Bureau Young Research Advisors, National Children’ Bureau Family Research Advisory Groups, NIHR Great Ormond Street Hospital (GOSH) Biomedical Research Centre (BRC) Parent and Carer Advisory Group, GOSH Young People’s Forum and GOSH Young Persons Advisory Group for their input to this project. We also gratefully acknowledge all children and families whose de-identified data are used in this analysis.

We would like to thank Nicolas Libuy, Pia Hardelid, Chloe Parkin and Matthew Lilliman for their contributions to this project.

## Ethics approval

Approvals for the use of HES data were obtained as part of the standard Hospital Episode Statistics approval process and ethical approval was obtained from London—South East Research Ethics Committee (reference 16/LO/0012). HES records were made available by NHS Digital.

## Authors’ contributions

The study was conceived by RG, KH, and RB. DE conducted all the analyses with input from KH and RG. DE wrote the manuscript with input from all the authors. All the authors approved the final manuscript.

## Data availability

The data used in this analysis is expected to be available to accredited researchers in 2022 (as part of the ECHILD Database) by applying to the data providers (DfE and NHS Digital).

