## Supplemental material for "Deficits in hospital care among clinically vulnerable children aged 0 to 4 years during the COVID-19 pandemic"

**Supplementary Table 1: Average percentage of 0–4-year-olds with at least one hospital contact per year (2015-19), by risk factors.**

| | | | 2015-19 averages (%) | | | | | |
|---|---|---|---|---|---|---|---|---|
| | | n (%) | Outpatient attendances | p-value | Planned admissions | p-value | Unplanned admissions | p-value |
| **Clinical vulnerability** | No known vulnerability | 2,333,773 (72.2) | 14.1 | Ref | 1.74 | Ref | 8.8 | Ref |
| | Any vulnerability | 644,933 (20.0) | 31.2 | <0.001 | 10.3 | <0.001 | 18.4 | <0.001 |
| | • LBW/Preterm only | 238,559 (7.4) | 17.8 | <0.001 | 1.65 | 0.012 | 8.4 | <0.001 |
| | • CC only | 329,072 (10.2) | 39.1 | <0.001 | 16.0 | <0.001 | 25.3 | <0.001 |
| | • Both | 77,302 (2.4) | 38.8 | <0.001 | 12.3 | <0.001 | 20.0 | <0.001 |
| | Any CC$^{\alpha}$ | 406,374 (12.6) | 38.9 | <0.001 | 15.3 | <0.001 | 24.3 | <0.001 |
| **Age group** | Infants (<1 year old) | 628,900 (19.5) | 60.1 | <0.001 | 4.2 | <0.001 | 26.4 | <0.001 |
| | 1–4-year-olds | 2,601,649 (80.5) | 8.2 | Ref | 3.5 | Ref | 8.0 | Ref |
| **Ethnic group** | White | 2,296,986 (71.1) | 17.7 | Ref | 3.7 | Ref | 12.0 | Ref |
| | Black | 153,009 (4.7) | 15.8 | <0.001 | 3.8 | 0.018 | 9.2 | <0.001 |
| | Asian | 345,843 (10.7) | 18.0 | <0.001 | 3.6 | 0.007 | 11.6 | <0.001 |
| | Other | 257,651 (8.0) | 19.1 | <0.001 | 3.6 | 0.006 | 10.7 | <0.001 |
| **Deprivation quintile** | Q1 (most deprived) | 755,351 (23.4) | 19.5 | <0.001 | 4.4 | 0.304 | 14.0 | <0.001 |
| | Q2 | 607,815 (18.8) | 19.6 | <0.001 | 4.2 | 0.003 | 13.3 | <0.001 |
| | Q3 | 502,672 (15.6) | 20.2 | <0.001 | 4.2 | <0.001 | 13.5 | 0.071 |
| | Q4 | 430,486 (13.3) | 21.2 | <0.001 | 4.3 | 0.086 | 13.8 | 0.12 |
| | Q5 (least deprived) | 404,753 (12.5) | 22.5 | Ref | 4.3 | Ref | 13.6 | Ref |

**LBW** – Low birthweight; **CC** – Chronic conditions. * N = 3,230,549 (Average eligible population between 2015-2019). Ref – Reference group. P-values: Chi-square test for comparison with reference group. α Children with any chronic condition (combines CC only and Both).

**Supplementary Table 2: Difference in predicted and observed rates of hospital contact during the pandemic (March 2020-2021) among children aged 0 to 4 years, by age.**

| | Percentage of children seen | | | Number of hospital contacts | | | | Rates per 1000 child-years | | | |
|---|---|---|---|---|---|---|---|---|---|---|---|
| | n | N | % | Predicted | Observed | Difference | % change | Predicted | Observed | Difference (95% CI) | % change |
| **Outpatient attendances** | | | | | | | | | | | |
| **Total** | 426,871 | 3,034,197 | 14.1 | 1,687,100 | 1,282,399 | -404,701 | -24.0 | 556 | 423 | **-133** (-131, -136) | -24.0 |
| Infants (<1 year old) | 313,236 | 568,812 | 55.1 | 900993 | 815513 | -85,480 | -9.5 | 1584 | 1434 | **-150*** (-143, -157) | -9.5 |
| 1–4-year-olds | 113,635 | 2,465,385 | 4.6 | 786107 | 466886 | -319,221 | -40.6 | 319 | 189 | **-129** (-128, -131) | -40.6 |
| **Planned admissions** | | | | | | | | | | | |
| **Total** | 63,439 | 3,034,197 | 2.1 | 166,954 | 104,464 | -62,490 | -37.4 | 55 | 34 | **-21** (-20, -21) | -37.4 |
| Infants (<1 year old) | 19,597 | 568,812 | 3.4 | 35045 | 28108 | -6,937 | -19.8 | 62 | 49 | **-12*** (-11, -14) | -19.8 |
| 1–4-year-olds | 43,842 | 2,465,385 | 1.8 | 131909 | 76356 | -55,553 | -42.1 | 54 | 31 | **-23** (-22, -23) | -42.1 |
| **Unplanned admissions** | | | | | | | | | | | |
| **Total** | 208,377 | 3,034,197 | 6.9 | 519,640 | 268,220 | -251,420 | -48.4 | 171 | 88 | **-83** (-81, -84) | -48.4 |
| Infants (<1 year old) | 106,048 | 568,812 | 18.6 | 226396 | 137884 | -88,512 | -39.1 | 398 | 242 | **-156*** (-152, -159) | -39.1 |
| 1–4-year-olds | 102,329 | 2,465,385 | 4.2 | 293244 | 130336 | -162,908 | -55.6 | 119 | 53 | **-66** (-65, -67) | -55.6 |

* Significantly different from 1–4-year-olds (5% level of significance).

**Supplementary Table 3: Difference in predicted and observed rates of hospital contact during the pandemic (March 2020-2021) among children aged 0 to 4 years, by ethnic group.**

| | Percentage of children seen | | | Number of hospital contacts | | | | Rates per 1000 child-years | | | |
|---|---|---|---|---|---|---|---|---|---|---|---|
| | n | N | % | Predicted | Observed | Difference | % change | Predicted | Observed | Difference (95% CI) | % change |
| **Outpatient attendances** | | | | | | | | | | | |
| **Total** | 380,925 | 2,872,476 | 13.3 | 1,589,161 | 1,203,043 | -386,118 | -24.3 | 553 | 419 | **-134** (-131, -138) | -24.3 |
| White | 277,757 | 2,125,238 | 13.1 | 1177279 | 889991 | -287,288 | -24.4 | 554 | 419 | **-135** (-133, -137) | -24.4 |
| Black | 17,582 | 140,621 | 12.5 | 70897 | 52346 | -18,551 | -26.2 | 504 | 372 | **-132** (-124, -140) | -26.2 |
| Asian | 48,025 | 338,198 | 14.2 | 195379 | 150173 | -45,206 | -23.1 | 578 | 444 | **-134** (-128, -139) | -23.1 |
| Other | 37,561 | 268,419 | 14.0 | 145606 | 110533 | -35,073 | -24.1 | 542 | 412 | **-131** (-125, -137) | -24.1 |
| **Planned admissions** | | | | | | | | | | | |
| **Total** | 60,815 | 2,872,476 | 2.1 | 161,107 | 102,311 | -58,796 | -36.5 | 56 | 36 | **-20** (-19, -22) | -36.5 |
| White | 45,439 | 2,125,238 | 2.1 | 116675 | 75721 | -40,954 | -35.1 | 55 | 36 | **-19** (-19, -20) | -35.1 |
| Black | 3,053 | 140,621 | 2.2 | 8443 | 4798 | -3,645 | -43.2 | 60 | 34 | **-26*** (-23, -29) | -43.2 |
| Asian | 6,790 | 338,198 | 2.0 | 20335 | 12319 | -8,016 | -39.4 | 60 | 36 | **-24*** (-22, -26) | -39.4 |
| Other | 5,533 | 268,419 | 2.1 | 15654 | 9473 | -6,181 | -39.5 | 58 | 35 | **-23*** (-21, -25) | -39.5 |
| **Unplanned admissions** | | | | | | | | | | | |
| **Total** | 199,183 | 2,872,476 | 6.9 | 498,620 | 260,250 | -238,370 | -47.8 | 174 | 91 | **-83** (-81, -85) | -47.8 |
| White | 152,225 | 2,125,238 | 7.2 | 378452 | 199614 | -178,838 | -47.3 | 178 | 94 | **-84** (-83, -85) | -47.3 |
| Black | 7,566 | 140,621 | 5.4 | 18489 | 9442 | -9,047 | -48.9 | 131 | 67 | **-64*** (-60, -69) | -48.9 |
| Asian | 22,516 | 338,198 | 6.7 | 61164 | 29575 | -31,589 | -51.6 | 181 | 87 | **-93*** (-90, -97) | -51.6 |
| Other | 16,876 | 268,419 | 6.3 | 40515 | 21619 | -18,896 | -46.6 | 151 | 81 | **-70*** (-67, -74) | -46.6 |

* Significantly different from children in the white ethnic group (5% level of significance). Other ethnic group includes Chinese and mixed ethnicity.

**Supplementary Table 4: Difference in predicted and observed rates of hospital contact during the pandemic (March 2020-2021) among children aged 0 to 4 years, by deprivation quintile.**

| | Percentage of children seen | | | Number of hospital contacts | | | | Rates per 1000 child-years | | | |
|---|---|---|---|---|---|---|---|---|---|---|---|
| | n | N | % | Predicted | Observed | Difference | % change | Predicted | Observed | Difference (95% CI) | % change |
| **Outpatient attendances** | | | | | | | | | | | |
| **Total** | 389,098 | 2,733,692 | 14.2 | 1,433,837 | 1,213,380 | -220,457 | -15.4 | 525 | 444 | **-81** (-78, -86) | -15.4 |
| Q1 | 103,866 | 750,360 | 13.8 | 405740 | 331783 | -73,957 | -18.2 | 541 | 442 | **-99*** (-95, -102) | -18.2 |
| Q2 | 84,205 | 604,305 | 13.9 | 304926 | 257812 | -47,114 | -15.5 | 505 | 427 | **-78** (-74, -82) | -15.5 |
| Q3 | 72,458 | 512,075 | 14.1 | 260352 | 226099 | -34,253 | -13.2 | 508 | 442 | **-67*** (-63, -71) | -13.2 |
| Q4 | 65,403 | 447,272 | 14.6 | 234302 | 202578 | -31,724 | -13.5 | 524 | 453 | **-71** (-67, -75) | -13.5 |
| Q5 (least deprived) | 63,166 | 419,680 | 15.1 | 228517 | 195108 | -33,409 | -14.6 | 545 | 465 | **-80** (-75, -84) | -14.6 |
| **Planned admissions** | | | | | | | | | | | |
| **Total** | 62,744 | 2,733,692 | 2.3 | 146,454 | 104,255 | -42,199 | -28.8 | 54 | 38 | **-16** (-14, -17) | -28.8 |
| Q1 | 17,518 | 750,360 | 2.3 | 42046 | 28414 | -13,632 | -32.4 | 56 | 38 | **-18*** (-17, -19) | -32.4 |
| Q2 | 13,703 | 604,305 | 2.3 | 32566 | 22934 | -9,632 | -29.6 | 54 | 38 | **-16*** (-15, -17) | -29.6 |
| Q3 | 11,625 | 512,075 | 2.3 | 26400 | 19069 | -7,331 | -27.8 | 52 | 37 | **-14** (-13, -16) | -27.8 |
| Q4 | 10,163 | 447,272 | 2.3 | 23504 | 17253 | -6,251 | -26.6 | 53 | 39 | **-14** (-13, -15) | -26.6 |
| Q5 (least deprived) | 9,735 | 419,680 | 2.3 | 21938 | 16585 | -5,353 | -24.4 | 52 | 40 | **-13** (-11, -14) | -24.4 |
| **Unplanned admissions** | | | | | | | | | | | |
| **Total** | 206,549 | 2,733,692 | 7.6 | 459,218 | 267,418 | -191,800 | -41.8 | 168 | 98 | **-70** (-68, -73) | -41.8 |
| Q1 | 57,328 | 750,360 | 7.6 | 133406 | 74919 | -58,487 | -43.8 | 178 | 100 | **-78*** (-76, -80) | -43.8 |
| Q2 | 44,184 | 604,305 | 7.3 | 98003 | 57071 | -40,932 | -41.8 | 162 | 94 | **-68** (-66, -70) | -41.8 |
| Q3 | 38,374 | 512,075 | 7.5 | 84028 | 49672 | -34,356 | -40.9 | 164 | 97 | **-67** (-65, -69) | -40.9 |
| Q4 | 34,434 | 447,272 | 7.7 | 74938 | 44550 | -30,388 | -40.6 | 168 | 100 | **-68** (-65, -71) | -40.6 |
| Q5 (least deprived) | 32,229 | 419,680 | 7.7 | 68843 | 41206 | -27,637 | -40.1 | 164 | 98 | **-66** (-63, -68) | -40.1 |

* Significantly different from children in the least deprived quintile (5% level of significance).

**Supplementary Table 5: Difference in predicted and observed rates of hospital contact per 1,000 child-years among children aged 0 to 4 years during the pandemic (March 2020-2021), by period and age.**

| | March 23 to June 23, 2020 | | June 24 to Nov 4, 2020 | | Nov 5 to Dec 31, 2020 | | Jan 1 to March 22, 2021 | |
|---|---|---|---|---|---|---|---|---|
| | Deficit (95% CI) | % change | Deficit (95% CI) | % change | Deficit (95% CI) | % change | Deficit (95% CI) | % change |
| **Outpatient attendances** | | | | | | | | |
| **Total** | **-170** (-167, -172) | -30.7 | **-90** (-88, -92) | -16.6 | **-101** (-98, -104) | -18.8 | **-184** (-181, -188) | -30.7 |
| Infants (<1 year old) | **-310*** (-304, -317) | -20.2 | **-37*** (-32, -43) | -2.4 | **-70*** (-62, -79) | -4.5 | **-207*** (-198, -217) | -12.3 |
| 1–4-year-olds | **-137** (-136, -139) | -43.9 | **-102** (-101, -103) | -32.9 | **-108** (-106, -110) | -36.0 | **-179** (-177, -181) | -51.3 |
| **Planned admissions** | | | | | | | | |
| **Total** | **-33** (-32, -33) | -60.1 | **-16** (-15, -17) | -29.1 | **-9** (-8, -10) | -17.6 | **-22** (-21, -23) | -38.2 |
| Infants (<1 year old) | **-25*** (-23, -26) | -40.8 | **-8*** (-7, -9) | -13.1 | **-2*** (0, -4) | -3.3 | **-12*** (-10, -14) | -18.3 |
| 1–4-year-olds | **-35** (-34, -35) | -65.2 | **-18** (-17, -18) | -33.4 | **-10** (-10, -11) | -21.6 | **-24** (-24, -25) | -43.4 |
| **Unplanned admissions** | | | | | | | | |
| **Total** | **-82** (-81, -83) | -53.2 | **-60** (-59, -61) | -38.2 | **-124** (-122, -126) | -55.5 | **-92** (-91, -94) | -52.1 |
| Infants (<1 year old) | **-150*** (-147, -153) | -41.4 | **-103*** (-100, -106) | -28.6 | **-288*** (-283, -293) | -53.4 | **-154*** (-149, -159) | -38.7 |
| 1–4-year-olds | **-66** (-65, -67) | -62.6 | **-50** (-49, -50) | -45.4 | **-86** (-85, -87) | -57.2 | **-78** (-77, -80) | -61.8 |

* Significantly different from 1–4-year-olds (5% level of significance).

**Supplementary Table 6: Difference in predicted and observed rates of hospital contact per 1,000 child-years among children aged 0 to 4 years during the pandemic (March 2020-2021), by period and ethnic group.**

| | March 23 to June 23, 2020 | | June 24 to Nov 4, 2020 | | Nov 5 to Dec 31, 2020 | | Jan 1 to March 22, 2021 | |
|---|---|---|---|---|---|---|---|---|
| | Deficit (95% CI) | % change | Deficit (95% CI) | % change | Deficit (95% CI) | % change | Deficit (95% CI) | % change |
| **Outpatient attendances** | | | | | | | | |
| **Total** | **-178** (-175, -181) | -34.1 | **-99** (-97, -102) | -18.1 | **-111** (-107, -115) | -20.5 | **-157** (-153, -161) | -27.7 |
| White | **-179** (-177, -181) | -32.3 | **-99** (-97, -101) | -18.0 | **-118** (-116, -121) | -21.7 | **-155** (-153, -158) | -27.6 |
| Black | **-178** (-170, -185) | -36.1 | **-101** (-94, -107) | -20.1 | **-67*** (-58, -76) | -14.1 | **-175*** (-164, -186) | -32.5 |
| Asian | **-179** (-174, -185) | -31.5 | **-103** (-98, -107) | -17.9 | **-87*** (-80, -93) | -15.8 | **-163** (-156, -171) | -26.9 |
| Other | **-176** (-170, -181) | -32.8 | **-98** (-93, -102) | -18.1 | **-105*** (-98, -112) | -19.9 | **-150** (-142, -158) | -26.8 |
| **Planned admissions** | | | | | | | | |
| **Total** | **-34** (-33, -35) | -60.1 | **-17** (-16, -18) | -29.4 | **-9** (-8, -10) | -17.7 | **-19** (-17, -20) | -33.5 |
| White | **-33** (-32, -33) | -59.2 | **-16** (-15, -16) | -27.8 | **-9** (-8, -10) | -17.3 | **-17** (-16, -18) | -31.4 |
| Black | **-37*** (-34, -39) | -62.4 | **-24*** (-22, -27) | -38.6 | **-11** (-8, -15) | -20.7 | **-26*** (-23, -30) | -44.0 |
| Asian | **-38*** (-36, -39) | -61.7 | **-20*** (-19, -22) | -33.5 | **-10** (-8, -12) | -17.4 | **-23*** (-21, -25) | -37.8 |
| Other | **-39*** (-37, -41) | -63.8 | **-18*** (-17, -20) | -30.7 | **-10** (-8, -12) | -18.9 | **-22*** (-19, -25) | -38.1 |
| **Unplanned admissions** | | | | | | | | |
| **Total** | **-85** (-83, -87) | -53.6 | **-62** (-61, -63) | -38.4 | **-128** (-125, -130) | -55.6 | **-83** (-81, -85) | -48.9 |
| White | **-84** (-83, -85) | -51.5 | **-63** (-62, -64) | -38.2 | **-135** (-134, -137) | -56.8 | **-82** (-81, -84) | -47.6 |
| Black | **-74*** (-70, -78) | -59.9 | **-48*** (-45, -51) | -37.9 | **-85*** (-80, -91) | -51.9 | **-65*** (-60, -71) | -52.1 |
| Asian | **-104*** (-101, -107) | -62.8 | **-67*** (-65, -70) | -40.4 | **-122*** (-118, -127) | -52.8 | **-102*** (-98, -107) | -55.9 |
| Other | **-77*** (-74, -80) | -55.5 | **-54*** (-51, -56) | -37.7 | **-95*** (-90, -99) | -49.5 | **-72*** (-68, -76) | -48.7 |

* Significantly different from children in the white ethnic group (5% level of significance).

**Supplementary Table 7: Difference in predicted and observed rates of hospital contact per 1,000 child-years among children aged 0 to 4 years during the pandemic (March 2020-2021), by period and deprivation quintile.**

| | March 23 to June 23, 2020 | | June 24 to Nov 4, 2020 | | Nov 5 to Dec 31, 2020 | | Jan 1 to March 22, 2021 | |
|---|---|---|---|---|---|---|---|---|
| | Deficit (95% CI) | % change | Deficit (95% CI) | % change | Deficit (95% CI) | % change | Deficit (95% CI) | % change |
| **Outpatient attendances** | | | | | | | | |
| **Total** | **-134** (-131, -138) | -25.4 | **-51** (-48, -54) | -9.6 | **-64** (-60, -69) | -12.4 | **-80** (-75, -85) | -15.5 |
| Q1 | **-151*** (-148, -155) | -28.0 | **-75*** (-72, -77) | -13.7 | **-74*** (-70, -78) | -13.9 | **-93** (-89, -98) | -17.6 |
| Q2 | **-126** (-123, -130) | -25.1 | **-50** (-47, -53) | -9.9 | **-64** (-60, -69) | -12.9 | **-77*** (-72, -81) | -15.5 |
| Q3 | **-126** (-122, -129) | -24.4 | **-34*** (-31, -37) | -6.7 | **-49*** (-44, -54) | -9.7 | **-65*** (-60, -70) | -13.1 |
| Q4 | **-127** (-123, -131) | -23.8 | **-35*** (-32, -39) | -6.7 | **-65** (-60, -70) | -12.5 | **-69*** (-64, -74) | -13.6 |
| Q5 (least deprived) | **-131** (-127, -136) | -23.9 | **-45** (-42, -49) | -8.3 | **-63** (-58, -69) | -11.9 | **-87** (-82, -93) | -16.3 |
| **Planned admissions** | | | | | | | | |
| **Total** | **-31** (-30, -32) | -56.0 | **-12** (-11, -13) | -21.8 | **-5** (-3, -6) | -9.2 | **-11** (-10, -13) | -21.8 |
| Q1 | **-33*** (-32, -35) | -58.2 | **-14*** (-13, -15) | -24.8 | **-7*** (-6, -9) | -14.0 | **-15*** (-13, -16) | -26.9 |
| Q2 | **-31*** (-30, -32) | -56.5 | **-13*** (-12, -14) | -22.8 | **-4** (-2, -5) | -7.6 | **-12*** (-11, -14) | -23.7 |
| Q3 | **-29** (-27, -30) | -55.1 | **-12** (-11, -13) | -22.3 | **-4** (-3, -6) | -8.6 | **-8** (-7, -10) | -17.7 |
| Q4 | **-30** (-28, -31) | -55.1 | **-10** (-9, -11) | -17.8 | **-3** (-1, -4) | -5.6 | **-11*** (-9, -12) | -21.7 |
| Q5 (least deprived) | **-28** (-27, -29) | -53.0 | **-10** (-9, -11) | -18.2 | **-3** (-2, -5) | -6.7 | **-6** (-5, -8) | -13.3 |
| **Unplanned admissions** | | | | | | | | |
| **Total** | **-76** (-74, -78) | -48.8 | **-51** (-49, -53) | -32.3 | **-116** (-113, -119) | -51.3 | **-62** (-60, -65) | -39.9 |
| Q1 | **-88*** (-86, -90) | -52.7 | **-56*** (-55, -58) | -33.9 | **-122*** (-119, 125) | -51.9 | **-70*** (-68, -73) | -42.0 |
| Q2 | **-77*** (-75, -79) | -50.2 | **-48** (-46, -49) | -31.7 | **-110** (-107, -113) | -50.3 | **-60** (-57, -62) | -40.0 |
| Q3 | **-71** (-69, -73) | -47.2 | **-47** (-45, -49) | -30.5 | **-116** (-112, -119) | -51.9 | **-61** (-58, -64) | -39.8 |
| Q4 | **-69** (-67, -71) | -45.2 | **-51** (-49, -53) | -32.2 | **-118** (-114, -121) | -51.8 | **-59** (-56, -62) | -37.9 |
| Q5 (least deprived) | **-67** (-65, -69) | -45.0 | **-51** (-49, -53) | -32.4 | **-110** (-107, -114) | -50.1 | **-57** (-54, -60) | -37.9 |

* Significantly different from children in the least deprived quintile (5% level of significance).

**Supplementary table 8: Type of outpatient attendance before and during the pandemic**

| | Pre-pandemic (Jan 1, 2015- Mar 22, 2020) | | During the pandemic (Mar 23, 2020- Mar 31, 2021) | |
|---|---|---|---|---|
| Outpatient attendance type | n | % | n | % |
| Attended in-person | 12,470,629 | 96.8 | 1,730,141 | 75.2 |
| Attended tele/virtual | 411,112 | 3.2 | 570,405 | 24.8 |
| Total | 12,261,444 | 100 | 2,300,546 | 100 |

**Supplementary Figure 1: Study population showing ascertainment of vulnerability status (exposure) from birth and hospital contacts (outcomes) from January 1, 2015 to March 31, 2021 for children aged 0 to 4 completed years. ***

| | | | | | | Outcome year | | | | | | |
|---|---|---|---|---|---|---|---|---|---|---|---|---|
| Year of birth | 2010 | 2011 | 2012 | 2013 | 2014 | 2015 | 2016 | 2017 | 2018 | 2019 | 2020 | 2021 |
| 2010 | <1 | 1 | 2 | 3 | 4 | 5 | 6 | 7 | 8 | 9 | 10 | 11 |
| 2011 | | <1 | 1 | 2 | 3 | 4 | 5 | 6 | 7 | 8 | 9 | 10 |
| 2012 | | | <1 | 1 | 2 | 3 | 4 | 5 | 6 | 7 | 8 | 9 |
| 2013 | | | | <1 | 1 | 2 | 3 | 4 | 5 | 6 | 7 | 8 |
| 2014 | | | | | <1 | 1 | 2 | 3 | 4 | 5 | 6 | 7 |
| 2015 | | | | | | <1 | 1 | 2 | 3 | 4 | 5 | 6 |
| 2016 | | | | | | | <1 | 1 | 2 | 3 | 4 | 5 |
| 2017 | | | | | | | | <1 | 1 | 2 | 3 | 4 |
| 2018 | | | | | | | | | <1 | 1 | 2 | 3 |
| 2019 | | | | | | | | | | <1 | 1 | 2 |
| 2020 | | | | | | | | | | | <1 | 1 |
| 2021 | | | | | | | | | | | | <1 |

**Exposure**

Low birth weight and preterm births

Chronic conditions

2 Age

**Outcomes**

Not considered

Partially considered

Fully considered

Not yet born

*Partially considered: For example, a child born on 1st June 2010 would turn 5 on 1st June 2015 and would therefore contribute data for part of 2015.

**Supplementary Figure 2: Pre-pandemic average rate of hospital contacts per 1,000 child-years among children aged 0 to 4 years (2015 to 2019), by clinical vulnerability status and risk factors.**

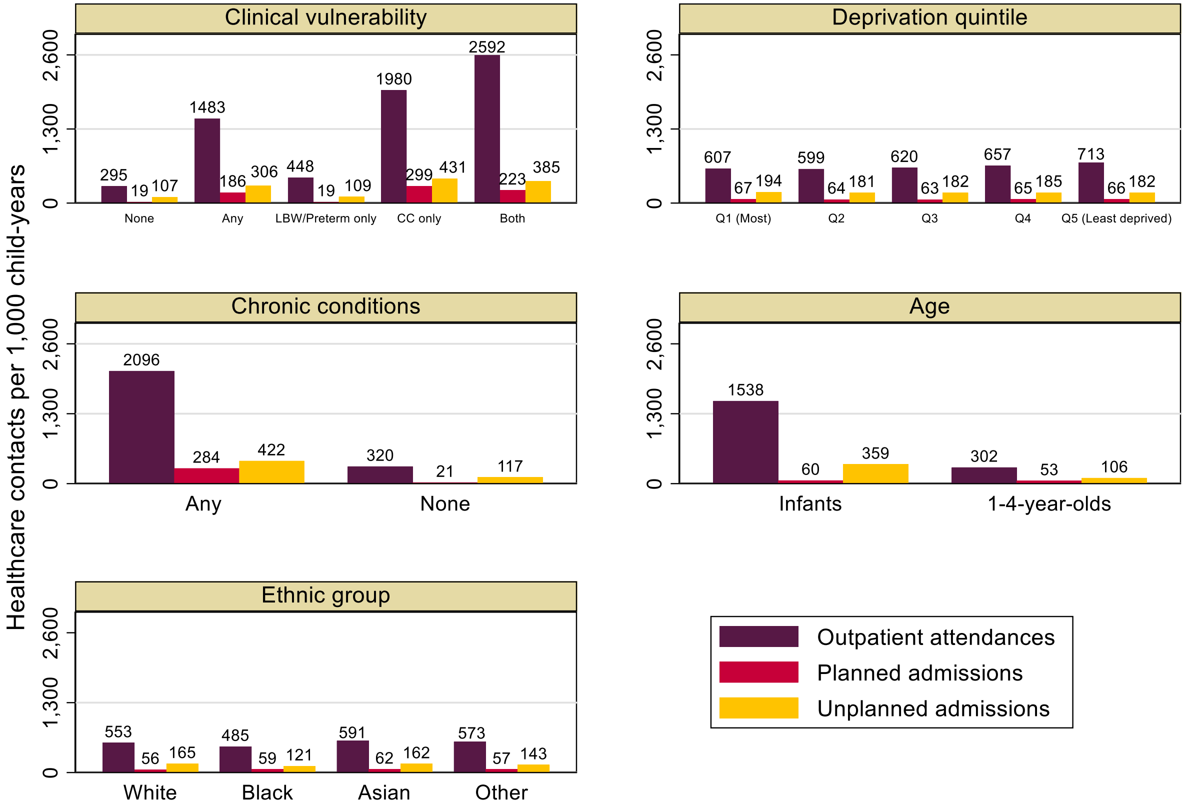


**LBW** – Low birthweight; **CC** – Chronic conditions

**Supplementary Figure 3: Pre-pandemic average rate of hospital contacts per 1,000 child-years among children aged 0 to 4 years (2015 to 2019), by ethnic group and presence of a chronic condition.**

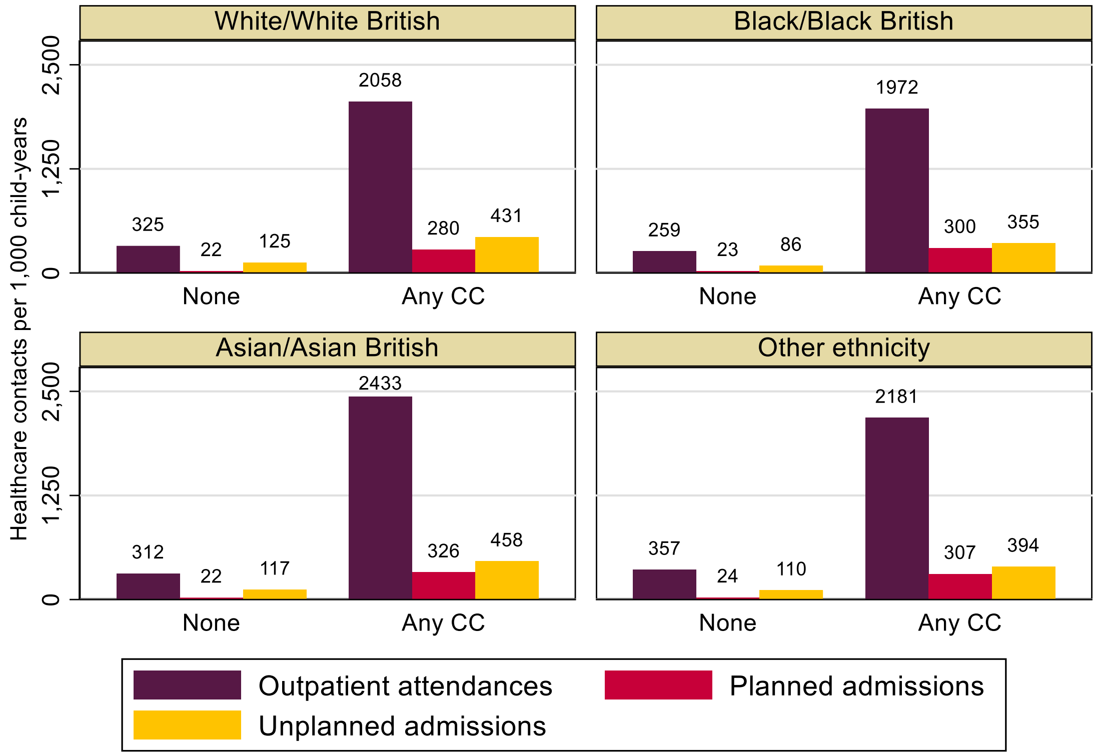


**CC** – Chronic conditions

**Supplementary Figure 4: Pre-pandemic average rate of hospital contacts per 1,000 child-years among children aged 0 to 4 years (2015 to 2019), by deprivation quintile and presence of a chronic condition.**

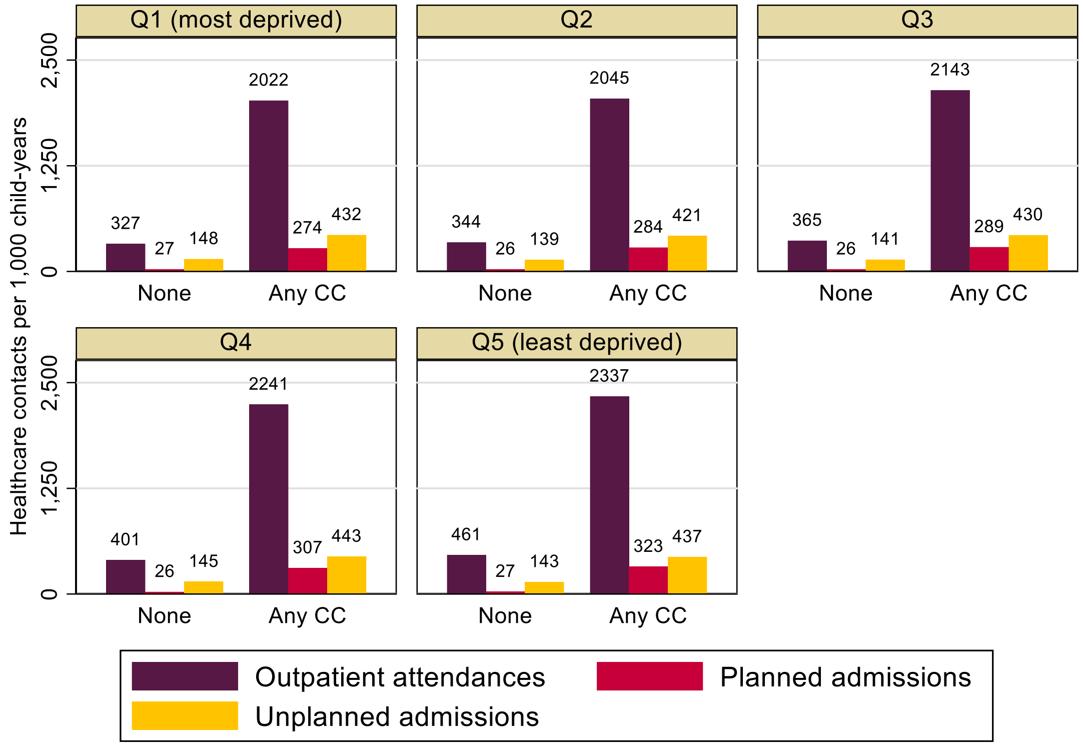


**CC** – Chronic conditions

**Supplementary Figure 5: Deficit in care during the pandemic (March 2020-2021), estimated from predicted minus observed rates of hospital contacts per 1,000 child-years for children aged 0 to 4 years, by ethnic group and presence of a chronic condition.**

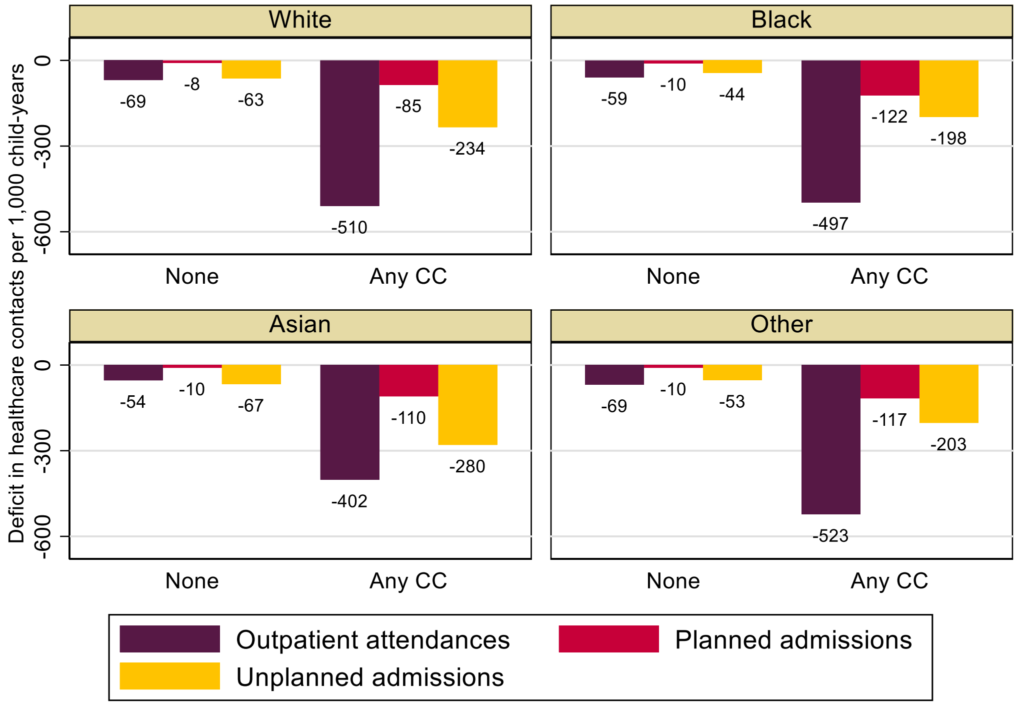


**CC** – Chronic conditions

**Supplementary Figure 6: Deficit in care during the pandemic (March 2020-2021), estimated from predicted minus observed rates of hospital contacts per 1,000 child-years for children aged 0 to 4 years, by deprivation quintile and presence of a chronic condition.**

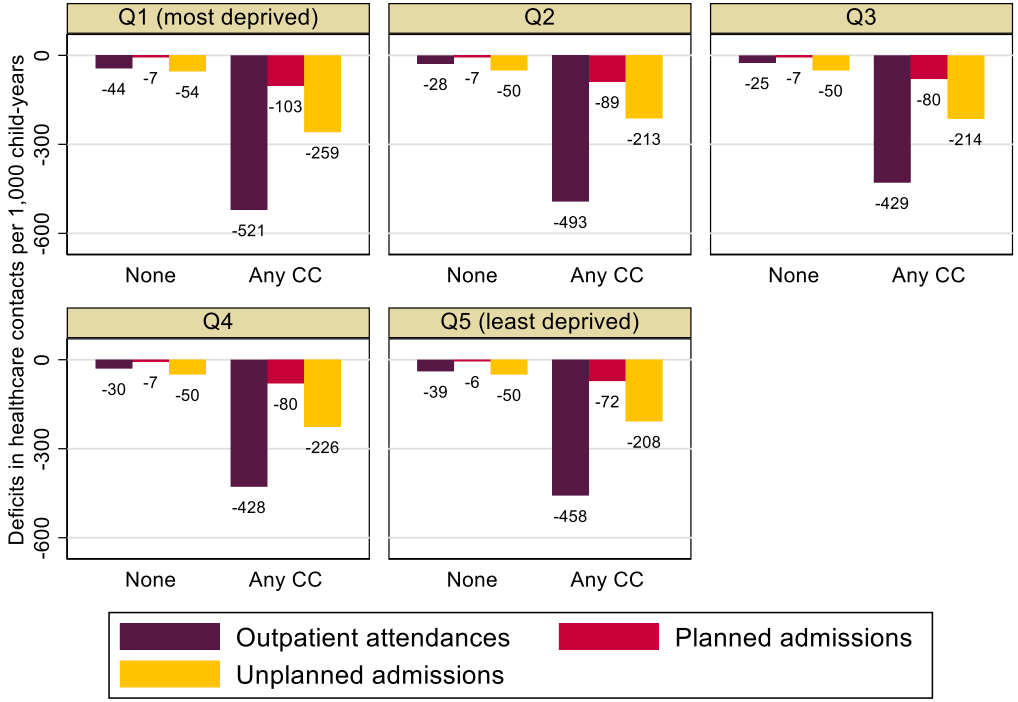


**CC** – Chronic conditions

**Supplementary Figure 7: Weekly difference in observed and predicted hospital contacts among children aged 0 to 4 years during the pandemic (March 2020-2021), by presence of a chronic condition.**

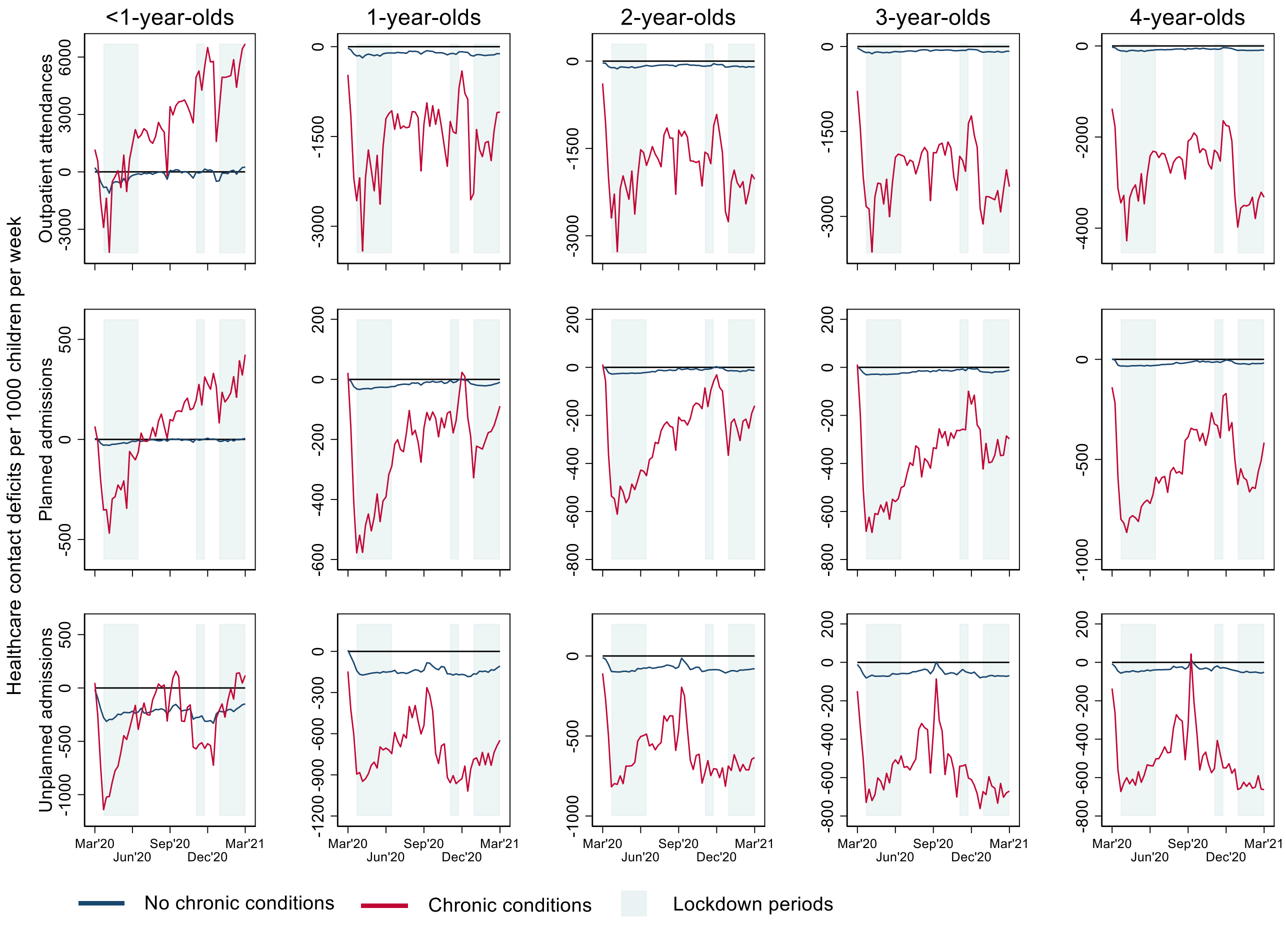

**Supplementary Figure 8: Weekly rates of hospital contacts among children aged 0 to 4 years during the pandemic (March 2020-2021) and on average from 2015-2019, by age comparing children in the most and least deprived IMD quintiles.**

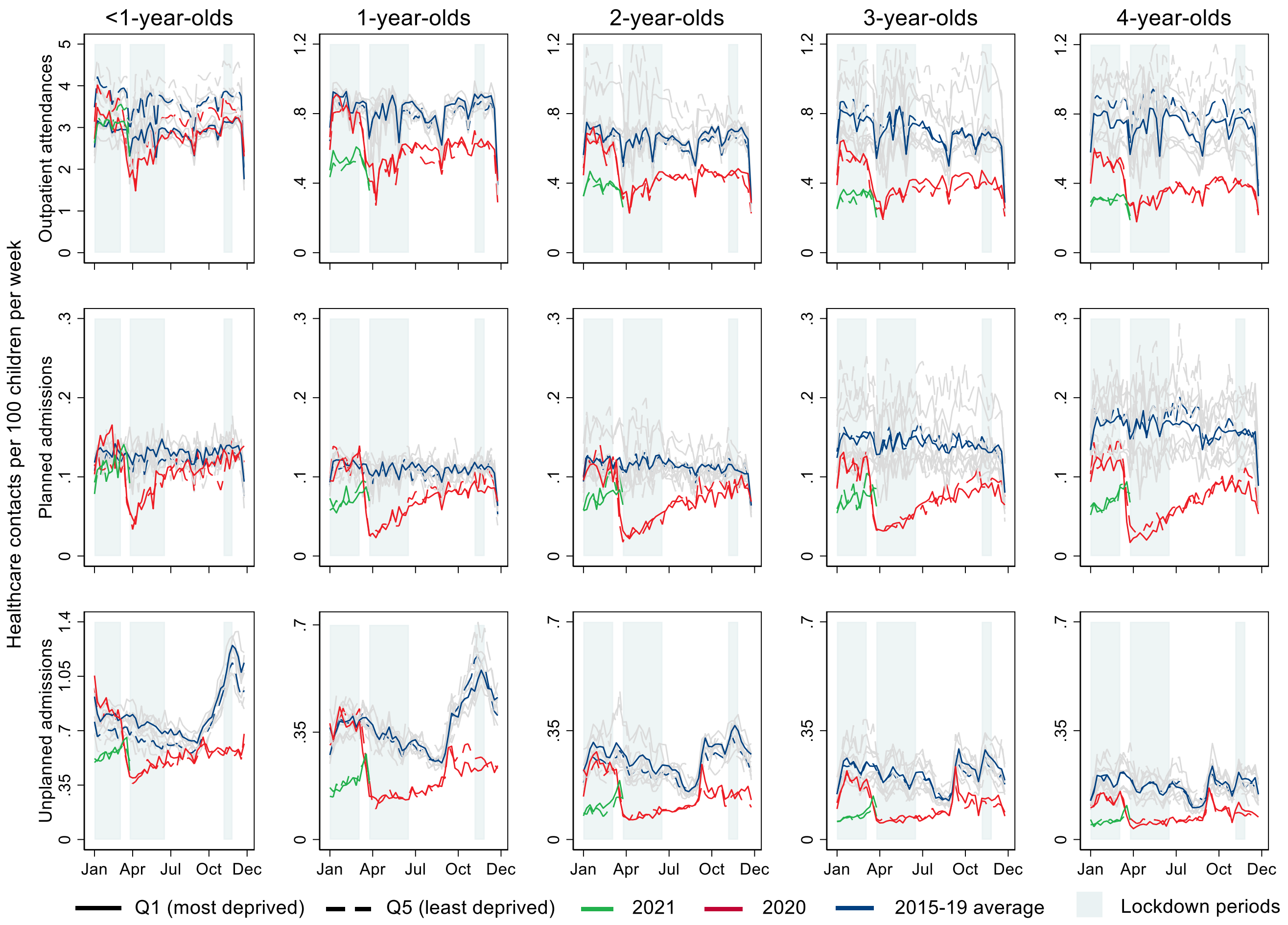


Note: January-March lockdown only affects 2021 data.

**Supplementary Figure 9: Weekly difference in observed and predicted hospital contacts among children aged 0 to 4 years during the pandemic (March 2020-2021), by deprivation quintile.**

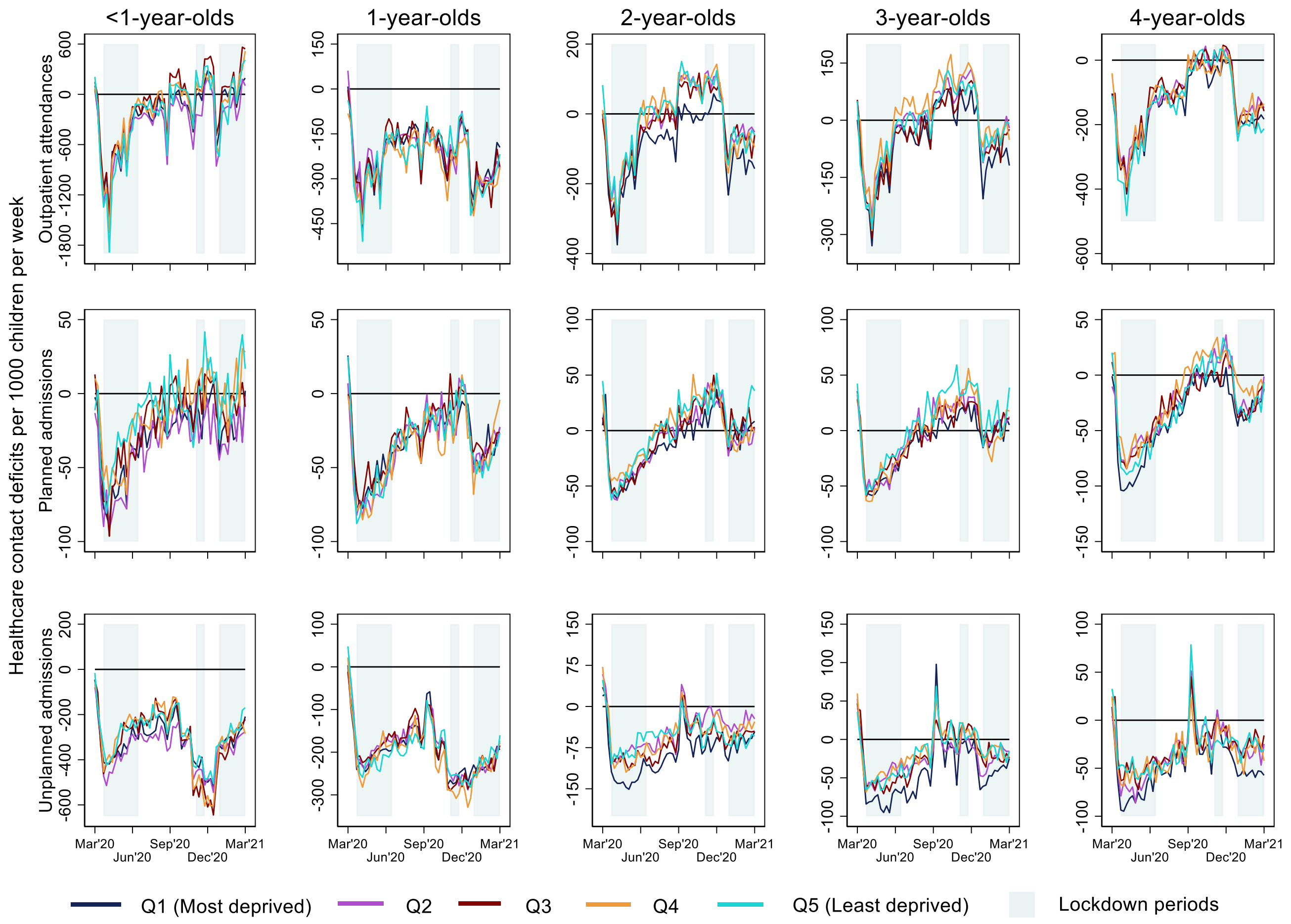

**Supplementary Figure 10: Weekly rates of hospital contacts among children aged 0 to 4 years during the pandemic (March 2020-2021) and on average from 2015-2019, by age comparing children of White and Asian ethnic groups.**

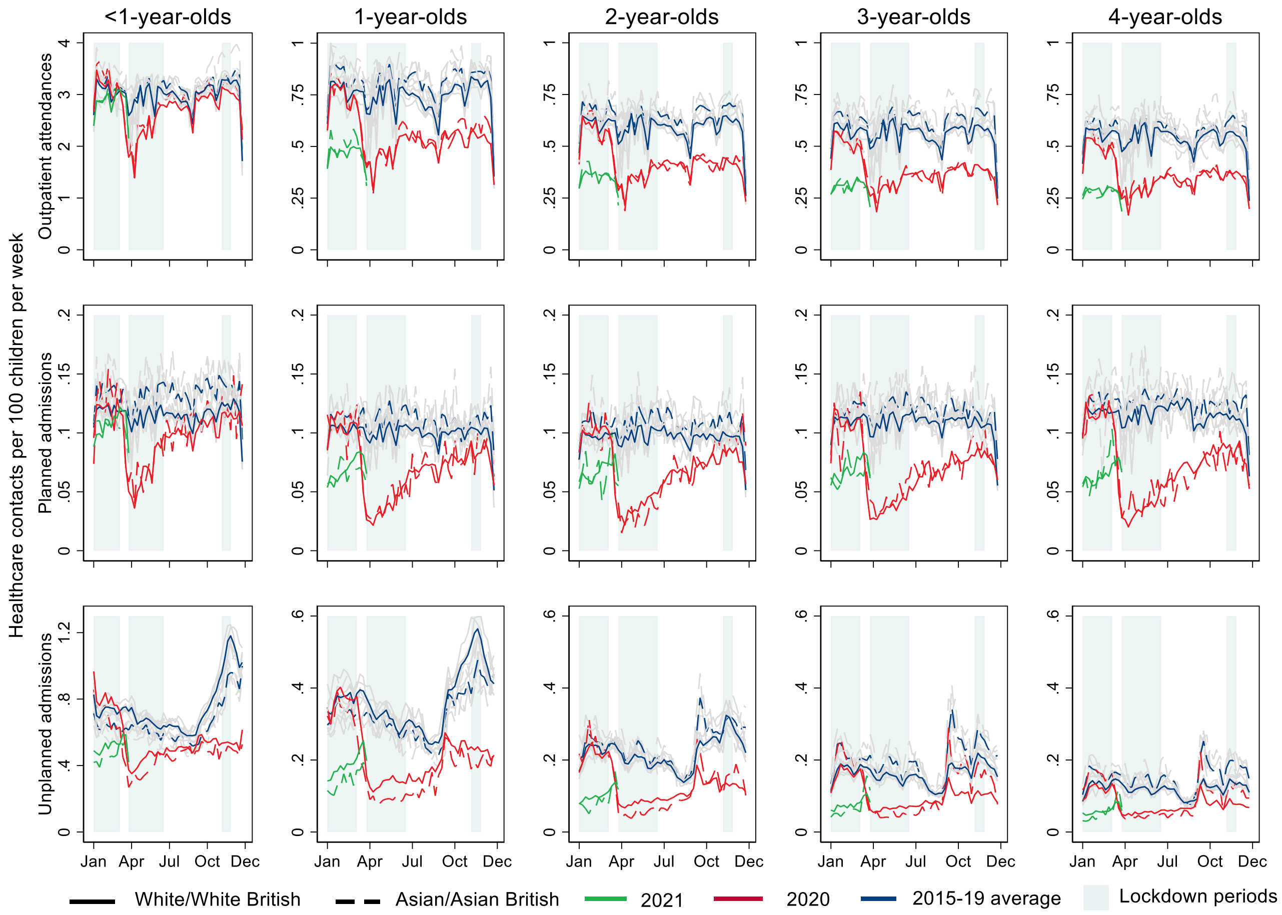


Note: January-March lockdown only affects 2021 data.

**Supplementary Figure 11: Weekly rates of hospital contacts among children aged 0 to 4 years during the pandemic (March 2020-2021) and on average from 2015-2019, by age comparing children of White and Black ethnic groups.**

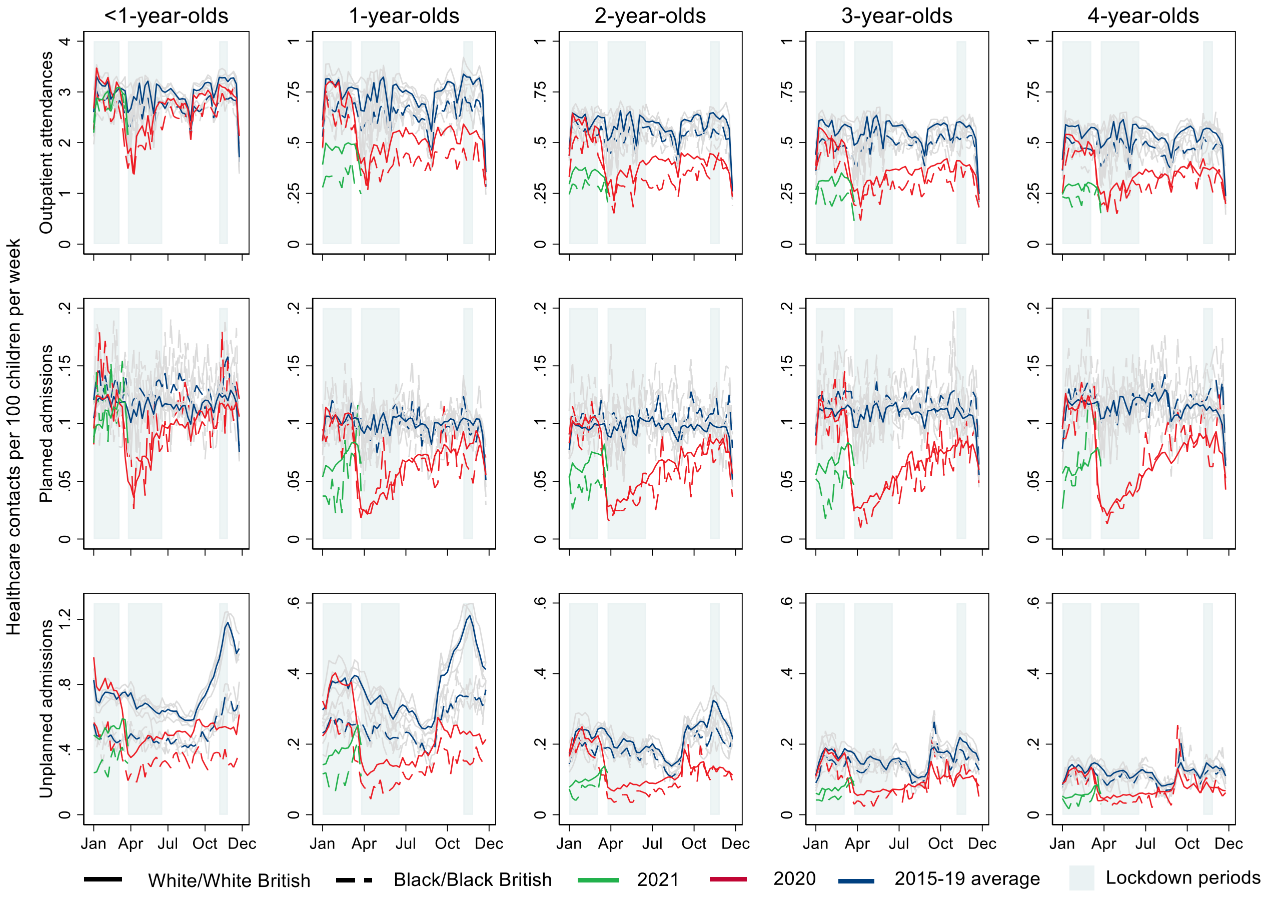


Note: January-March lockdown only affects 2021 data.

**Supplementary Figure 12: Weekly difference in observed and predicted hospital contacts among children aged 0 to 4 years during the pandemic (March 2020-2021), by ethnic group.**

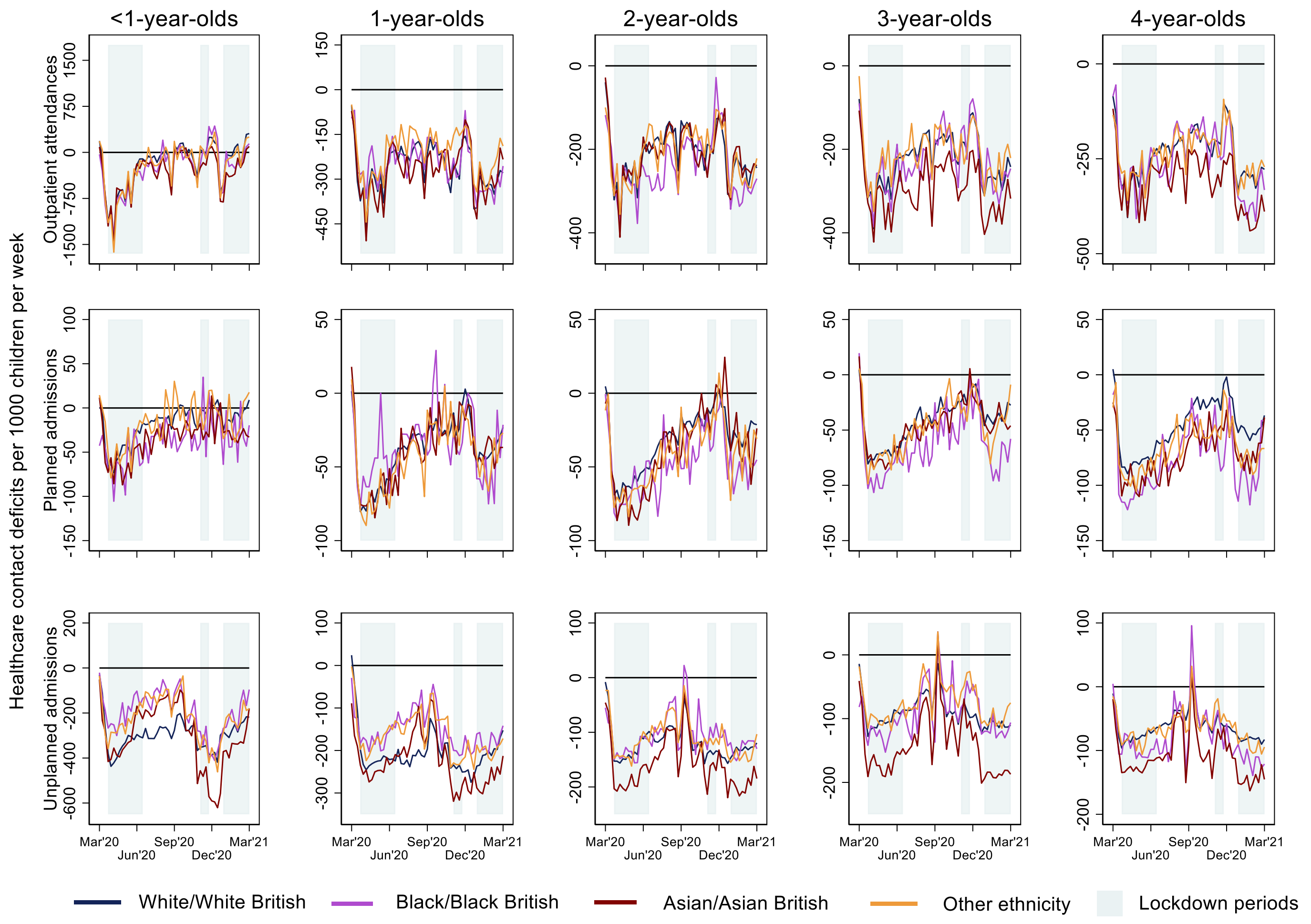

**Supplementary Figure 13: Weekly rates of in-person outpatient appointments among children aged 0 to 4 years during the pandemic (March 2020-2021) and on average from 2015-2019, by age.**

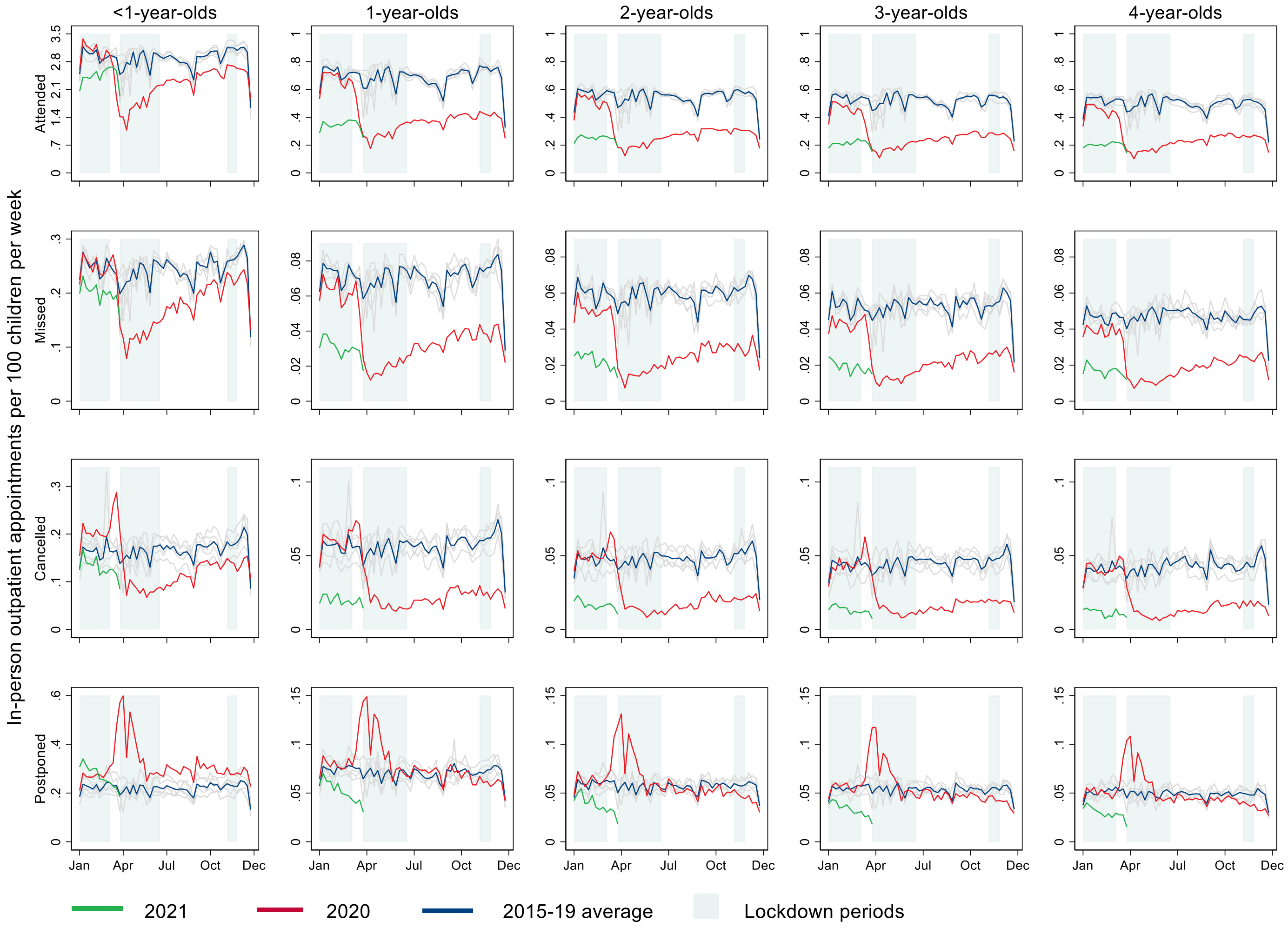


Note: January-March lockdown only affects 2021 data.

**Supplementary Figure 14: Weekly rates of outpatient attendances (In-person vs tele/virtual) among children aged 0 to 4 years during the pandemic (March 2020-2021) and on average from 2015-2019, by age.**

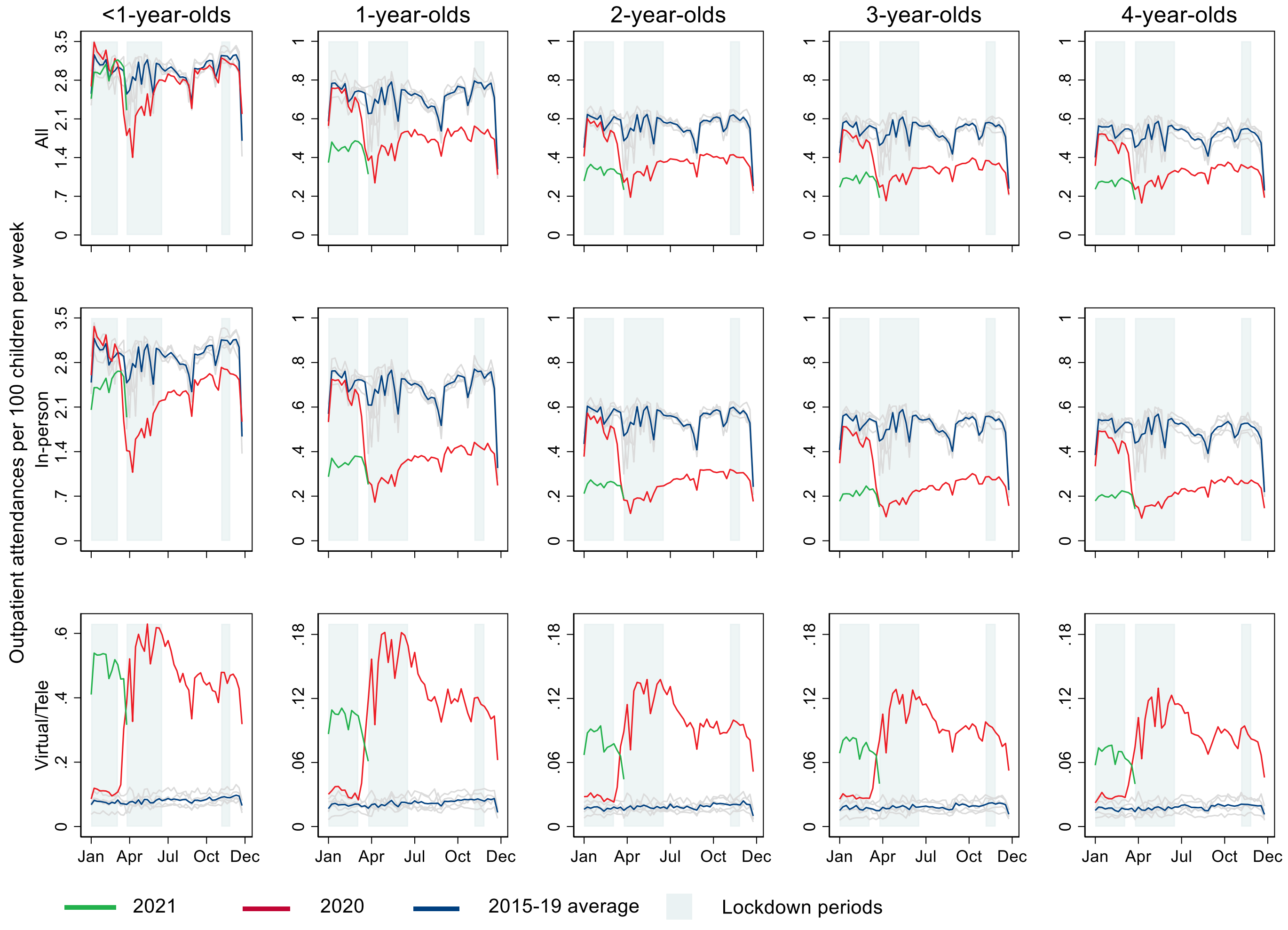


Note: January-March lockdown only affects 2021 data.
